# Highly efficient and low-cost single-cell culture platform for unbiased analysis of human memory B cell repertoire and antibody discovery

**DOI:** 10.1101/2025.01.14.25320505

**Authors:** Luciana Conde, Debora L. Oliveira, Gabriela Maciel, Fernando Castro, Aline de Oliveira Albuquerque, Danielle A.S. Rodrigues, Yare Mëllo, Gustavo Meira de Assis, Bárbara Gabrielle, Suyane S. Ferreira, Marcela S. Cunha, Carlena Navas, Manuela C. Emiliano, Marcele N. Rocha, Barbara Soares, Lucas Tostes, Philippe Caloba, Bruno Maia, Francisco M. Bastos de Oliveira, Amilcar Tanuri, Orlando C. Ferreira, Terezinha M.P.P. Castineiras, João Hermínio Martins da Silva, Juliana Echevarria, Marcelo Bozza, Leda R. Castilho, Luciana J. da Costa, Liza F. Felicori, Alberto Nobrega, Gabriel Victora, Carolina Lucas, Adriana Bonomo, André M. Vale

## Abstract

The SARS-CoV-2 pandemic underscored the need for innovative approaches to study humoral immunity and isolate monoclonal antibodies (mAbs) with diagnostic and therapeutic potential. Current methods for repertoire analysis at the clonal level require large-scale recombinant mAb production, limiting accessibility and delaying functional insight. We developed a single-cell culture (SCC) platform that enables profiling of human memory B cells (MBCs) and direct recovery of functional mAbs. Using samples from COVID-19 convalescent and vaccinated donors, we optimized SCCs with NB21 feeder cells, R848, and IL-2, achieving efficient clonal expansion and antibody secretion in short-term cultures. Screening and pseudovirus neutralization assays were performed directly with culture supernatants, bypassing the need for early recombinant antibody production. Antigen-baited cytometry sorting enriched Spike-specific MBCs by ∼30-fold. Among 592 isolated mAbs, 53% bound the Wuhan Spike, targeting the receptor-binding domain (28%), the N-terminal domain (15%), or other regions (57%). Cross-reactivity analysis revealed that 40% of anti-Spike mAbs recognized all tested variants of concern. VH/VL sequencing uncovered convergent rearrangements, including public V3-30 and V3-53/V3-66 clones, consistent with global findings. Two public RBD-specific antibodies demonstrated broad neutralization when produced recombinantly. Together, these results validate the SCC system as a streamlined approach for unbiased repertoire analysis and functional mAb isolation. More broadly, the platform provides a practical framework for linking B cell clonal composition with antigen specificity and serum antibody responses. By reducing costs and simplifying workflows, it expands opportunities for antibody discovery and immuno-epidemiological studies, fostering wider global participation in therapeutic antibody research.

## Introduction

The SARS-CoV-2 pandemic, with its high morbidity and mortality rates (1), highlighted the urgent need for new methods to study and monitor humoral immune responses to infection and vaccination at the level of individual B cell clonotypes. During the pandemic, immune escape was driven by the random accumulation of mutations in the viral genome, resulting in the emergence of highly transmissible Variants of Concern (VOCs) and significant antigenic drift in the Spike protein. These mutations, concentrated primarily in the receptor-binding domain (RBD) and the N-terminal domain (NTD), posed challenges to vaccination strategies and emphasized the need to monitor humoral responses in order to identify vaccine-induced cross-reactive and broadly neutralizing antibody responses (2).

Memory B cells (MBCs) are central to long-lasting immunity, generating high-affinity antibodies upon antigen re-encounter. These cells arise during primary antigen exposure and often undergo somatic hypermutation and affinity maturation in germinal centers, enhancing their ability to produce potent antibody responses (3). Upon reactivation, MBCs rapidly differentiate into antibody-secreting plasma cells, enabling efficient and robust secondary responses (4). Given their pivotal role in sustaining immune protection and their significant diversity, particularly against rapidly evolving pathogens, monitoring and analyzing MBC repertoires is a priority (5). This is particularly important for identifying monoclonal antibodies with therapeutic potential.

Single-cell high-throughput repertoire sequencing techniques have revolutionized the study of antibody responses, enabling detailed analyses of clonal evolution, somatic hypermutation, and lineage dynamics (6). These advancements have substantially enhanced our understanding of how diverse and effective antibody responses are generated and maintained. However, current high-throughput repertoire analysis is limited by their inability to simultaneously determine antigen-binding specificities alongside clonotypic variable Ig gene (VDJ/VJ) rearrangements. Presently, characterization of antigen specificity requires the cloning and expression of Ig genes carrying the identified VDJ/VJ rearrangements in cell lines to produce recombinant monoclonal antibodies (mAbs) for antigen-binding screening assays (7). This process is labor-intensive and time-consuming as it involves the production of hundreds recombinant mAbs to pinpoint the desired antibody. These technical demands are further compounded by financial barriers in many research settings, limiting widespread application of such approaches to pandemic and locally relevant pathogens.

*In vitro* culture methods of human B cells have been previously described as an alternative to the production of recombinant mAbs. A classical protocol described by Pinna et al. (8) requires co-cultures with peripheral blood mononuclear cells (PBMCs) from the same individual to act as feeder cells to support B cell growth. This approach allows the estimation of antigen-specific clonal frequencies and the study of the humoral response at the clonal level. While these cultures can efficiently stimulate single cell clonal expansion, they do not allow for unique recovery of VH/VL rearrangements from growing clones because of mRNA contamination from endogenous B cells in the PBMCs used as feeder cells.

In mouse experimental models, culture protocols employing feeder cell lines expressing activating B cell factors have been established as an efficient system for isolating monoclonal antibodies from naïve, and germinal center B cells (9–11). Similar culture systems have also been described for human B cells (12, 13); however, these protocols are generally more complex and delicate than those used in mice. They often require continuous supplementation with a cocktail of multiple cytokines, and the differentiation of antibody-secreting plasmablasts typically occurs only after 15–20 days in culture, much later than in murine systems.

Here, we describe a novel culture system for human B cell cultures that circumvents these limitations. By integrating elements from previously described single B cell culture protocol for both mouse and human, we developed a streamlined new approach based on the murine NB-21 feeder cell line, supplemented only with IL-2 and R848. This short-term (7-day) culture system yields sufficient amount of secreted Ig for antigen screening assays. It proved to be robust, reproducible, and highly efficient in supporting clonal expansion and Ig production, thereby establishing a platform for functional analysis of the human B cell repertoire. Our approach advances several key aspects of human monoclonal antibody discovery: (i) use of supernatants from short term single-cell cultured (SCCs) of memory B cells for antigen-specific screening, cross-reactivity studies across SARS-CoV-2 VOCs, and virus neutralization assays; (ii) analysis of VDJ/VJ gene usage from mRNA/cDNA derived directly from SCC lysates; (iii) high cloning efficiency that allows for an unbiased description of the antibody repertoire, and (iv) identification of mAbs candidates suitable for scalable recombinant production.

To validate the platform, we applied the SCC system to the study of the humoral response to SARS-CoV-2 infection. The results were consistent with previous landmark studies that employed high-throughput single-cell sequencing and recombinant mAbs production (14). Importantly, the protocol presented here offers a practical and accessible alternative for laboratories in low- and middle-income countries, enabling local health institutions to autonomously conduct critical immunological research and complementary epidemiological studies, as well as develop targeted monoclonal antibody therapies make significant contributions to clinical and global scientific efforts.

## Results

### Establishing culture conditions for human memory B cell expansion

Our research group pioneered the development of an efficient method for analyzing the mouse B cell repertoire, both for VH/VL rearrangements and antigen specificities, by combining high-efficiency B single-cell cultures (SCC) with RT-PCR for *Ig* variable gene amplification and sequencing (10). This approach relies on murine B cells cultured on S17 stromal cells (15), as feeder cells, in the presence of mitogens such as LPS, CpG, and Pam3Cys, to promote proliferation and differentiation into plasma cells *in vitro*. Following an analogous strategy, we designed a similar system for growing human memory B cells (MBCs) taking advantage of feeder cells able to provide essential factors for human B cell proliferation and antibody secretion.

To identify the most efficient feeder cell line for human MBC culture, we tested three previously described feeder cell lines: S17, 40LB, and NB21. S17, a mouse bone marrow stromal cell line which supports myelopoiesis and B lymphopoiesis. 40LB, derived from the 3T3 fibroblast lineage and engineered to express CD40L and BAFF, mimicking germinal center conditions (9). NB21, a derivative of 40LB, further engineered to express IL-21, enhancing mouse B cell survival and Ig secretion (16). To compare the performance of the different feeder cell types, sorted memory B cells (MBCs) from a healthy donor, seronegative donor for SARS-CoV-2 antibodies, were cultured in a limiting dilution assay (LDA) at varying cell densities, and the frequency of immunoglobulin-secreting cells were quantified after 7 days by ELISA. Applying the Poisson distribution, we calculated the frequency of clonable MBCs, defined as the seeding density at which 37% of cultures were negative for antibody detection. NB21 cells demonstrated significantly higher cloning efficiency, outperforming S17 and 40LB feeder lines by at least fourfold (**Supplementary Figures 1A, 1B**).

We confirmed that LPS was ineffective for stimulating human B cells, consistent with previous reports. Instead, optimal conditions for human MBC proliferation required R848 or CpG in combination with high doses of IL-2 (1,000 U/mL), as demonstrated by Pinna et al (2009) (8). Using NB21 as feeder cell, we compared R848 and CpG (both in the presence of hrIL-2) at concentrations of 2 μg/ml and 10nM, respectively. No significant differences in their efficacy to support memory B cell proliferation and antibody production were observed (**Supplementary Figure 1C**). Therefore, R848 was chosen for further experiments due to its cost-effectiveness and dual role as a TLR7 and TLR8 agonist (17).

Recognizing the need for methods adaptable to low-resource settings, we compared manual cell plating with automated robotic plating integrated into the sorting device. Both approaches yielded similar frequencies of responding B cells, with a representative experiment indicating 55% and 59% responder frequencies for robotic and manual plating, respectively (**Supplementary Figure 1D**).

### Validation of a culture system for comprehensive and unbiased MBC profiling

LDA using culture systems on efficient feeder cells enable reliable quantification of responding B cells (18, 19). We therefore applied this methodology to analyze the Spike-specific MBC repertoire in SARS-CoV-2-exposed donors and convalescent patients, with samples collected in 2020, before COVID-19 vaccination became available. SARS-CoV2-exposed donors were healthcare workers who had direct contact with patients, remained asymptomatic, and tested negative for SARS-CoV-2 by weekly PCR screening. These experiments aimed to assess the sensitivity and potential of our system for detecting and characterizing antigen-specific B cell responses.

We first assessed antibody titers of anti-SARS-CoV-2 Spike IgG in sera from two cohorts: (1) SARS-CoV2-exposed donors, and (2) convalescent patients from 2020. Additionally, as a control, we included 23 pre-COVID samples collected before 2019, prior to the emergence of SARS-CoV-2. Compared to pre-COVID samples, the exposed-negative group exhibited significantly higher levels of Spike-specific IgG. These findings suggest that the exposed-negative group may have developed some degree of antigen-specific immunity. This observation is consistent with prior report showing that individuals exposed to a virus do not necessarily develop viremia detectable by PCR, as observed among frontline healthcare professionals during the COVID-19 pandemic (20). Convalescent patients, sampled 20 to 180 days after symptom onset, exhibited higher IgG titers than the exposed-negative group (**Figure 1A**).

**Fig. 1:**
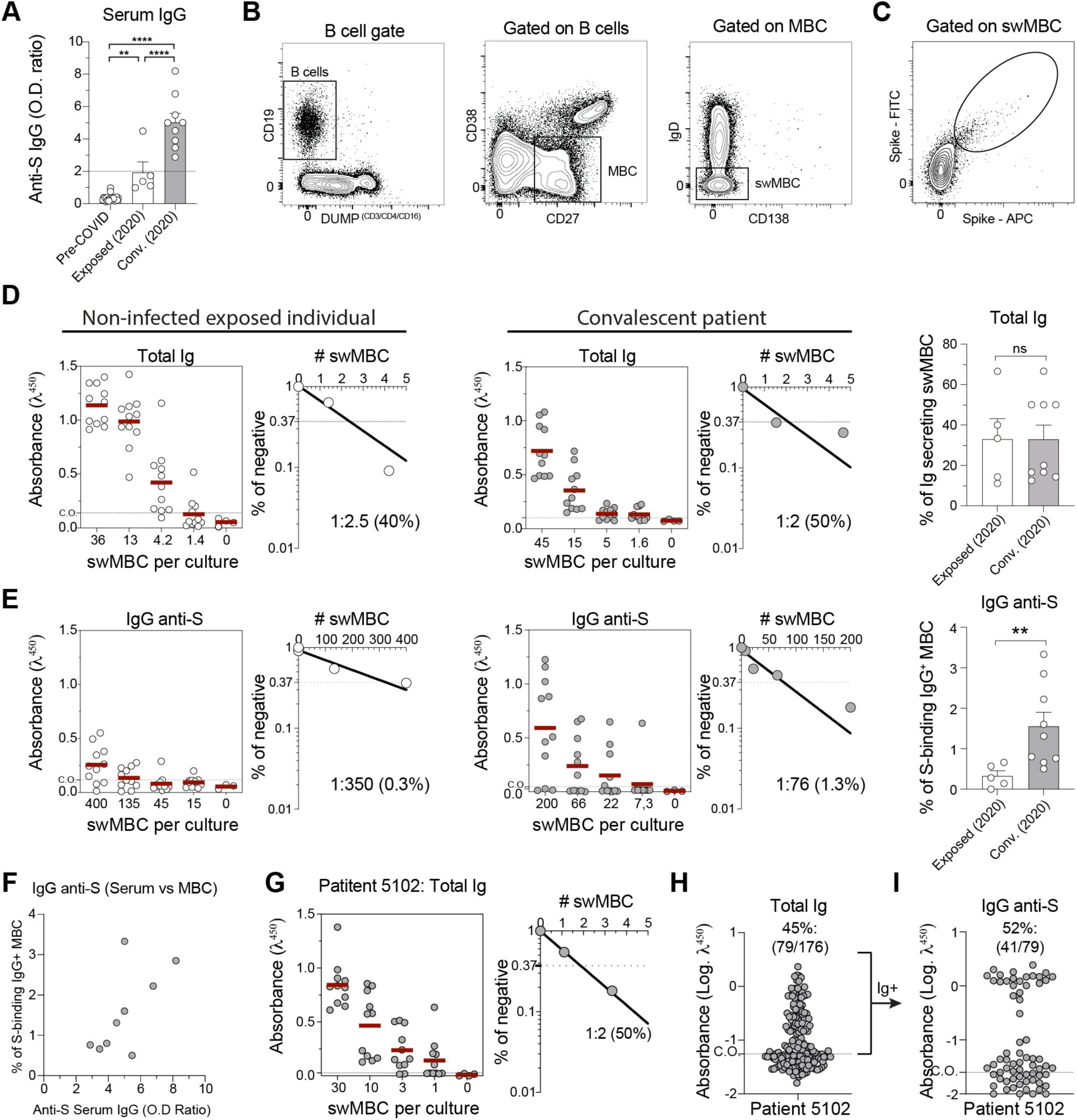
Validation of a culture method for comprehensive and unbiased analysis of the human memory B cell (MBC) repertoire. **(A)** Serological analysis showing the optical density (OD) ratio of Spike-specific IgG in pre-COVID, SARS-CoV-2-exposed but PCR-negative (“exposed-negative”), and convalescent patients, assayed by ELISA. Statistical analyses were performed using one-way ANOVA followed by Tukey’s multiple comparison test. **(B)** Gating strategy for sorting class switched MBCs (swMBCs). **(C)** Gating strategy for sorting Spike-baited swMBCs. **(D)** Limiting dilution assay (LDA) results from exposed-negative and convalescent patients, demonstrating the frequency of total Ig-secreting swMBC (representative donors shown on the left; pooled data shown in the dot plot). **(E)** Frequency of Spike-specific MBCs obtained from LDA culture of exposed-negative and convalescent patients (representative donors on the left; pooled data in the dot plot). Statistical analyses were performed using the unpaired two-tailed Student’s t test. **(F)** Correlation between the OD ratio of Spike-specific IgG in serum and the frequency of Spike-specific MBCs measured by LDA. **(G)** LDA results from a convalescent patient 5102, showing the frequency of total Ig secreting swMBC (50%). **(H)** SCC of Spike-baited swMBCs from donor 5102, showing 45% Ig-secreting cells, closely matching the frequency obtained by LDA. **(I)** Frequency of Spike-specific swMBCs from donor 5102 obtained in SCC (52% specific to Wuhan Spike). Significant difference is indicated by asterisk (*, P < 0.05; **, P < 0.01; (***, P < 0.001; ****, P < 0.0001). Error bars represent SEM.

The presence of anti-Spike antibodies in exposed-negative group prompted us to investigate whether our assay would be sensitive enough to detect relatively rare anti-S specific B cells within the MBC repertoire of SARS-CoV-2 in these individuals. To assess the frequency of Ig-secreting cells, we sorted total switched memory B cells (swMBCs) from both SARS-CoV-2 exposed-negative individuals and convalescent patients and cultured them under LDA conditions. The gating strategy used to isolate total swMBCs (CD19+CD27+CD38-IgD-) is shown in **Figure 1B**. For these initial experiments, we did not use the antigen-baited sorting strategy to enrich for S-specific MBCs as depicted in **Figure 1C**. Instead, total swMBCs were used to explore the unbiased repertoire. The percentage of Ig-secreting cells ranged from 14% to 60%, with no significant differences between negative and convalescent individuals (**Figure 1D**). Quantifying the number of Ig-secreting B cell clones per culture ensures accurate determination of the frequency of antigen-reactive B cells in each sample (8). To maintain an unbiased assessment of antigen-specific responses, we adjusted cell plating densities to calculate the percentage of B cells secreting Spike-specific antibodies. Exposed-negative individuals showed lower S-specific MBC frequencies (0.2%), compared to convalescent patients (1.6%) (**Figure 1E**). Interestingly, the frequency of IgG+ Spike-specific MBCs correlated with serum IgG anti-S levels in most convalescent donors (**Figure 1F**). These findings demonstrate that our culture method has sufficient sensitivity to identify anti-S B cell clones both in convalescent donors and exposed-negative individuals. LDA further revealed a ∼10-fold enrichment of anti-S clones in convalescent donors relative to exposed-negative individuals.

While LDA cultures can efficiently quantify antigen-specific MBCs frequencies, these cells represent only a small fraction of the overall MBC repertoire. Moreover, bulk culture LDA systems preclude mRNA isolation from individual B cell clones, impeding downstream transcriptional analysis of the antigen-specific VH/VL repertoire. To address this, we implemented single-cell cultures (SCCs) of antigen-baited sorted MBCs. In these experiments, S-baited MBCs were sorted using Wuhan strain Spike protein labeled with two different fluorochromes, APC and FITC, to ensure cell specificity to the antigen (**Figure 1C**). Spike-baited sorted cells from one patient (5102) were cultured individually with NB21 feeder cells and stimulated with R848 + IL-2. After 7 days, supernatants were analyzed for total Ig secretion and Spike-specific Ig by ELISA. The proportion of Ig-secreting cells in SCCs matched those observed in LDAs (**Figure 1G and H**), confirming that SCCs reliably support single B cell growth and differentiation. Of note, antigen-baiting strategy led to over 30-fold enrichment of Spike-specific cells, increasing their frequency from an average of 1.6% in LDA to ∼50% in SCC (**Figure 1G and H**). These results demonstrate that antigen-baited sorted MBCs can grow and differentiate under SCC conditions, facilitating the isolation of Spike-specific monoclonal antibodies and enabling downstream transcriptional analysis of VH/VL repertoires of individual antigen-specific clones.

### Single-cell cultures (SCCs) for antigen-directed Ig repertoire analysis and monoclonal antibody isolation

Our SCC system enables easy access to monoclonal antibodies in the culture supernatants of bait-selected MBCs for antigen-specific screening. To explore this capability, we analyzed a cohort of eight blood donors with a history of multiple SARS-CoV-2 vaccination and/or infection who exhibited high levels of serum-specific antibodies (**Supplementary Figure 2**). Samples were collected in 2022 and 2023, when the Omicron variant was the predominant strain. Spike-specific MBCs were sorted using a bait strategy and subjected to our SCC protocol. After 5 to 7 days of culture, approximately 250 µL of supernatant containing monoclonal Ig was obtained from each SCC well, providing sufficient volume to test each growing clone for total Ig secretion and multiple specificities of the secreted mAb by ELISA. For each individual donor, the total Ig secreted by individual MBC clones in the SCC are displayed in **Figure 2A**, with the frequency of responding B cells ranging from 22% to 54%. An average cloning efficiency of 37% across donors was observed, as measured by total Ig secretion (**Figure 2B**).

**Fig. 2:**
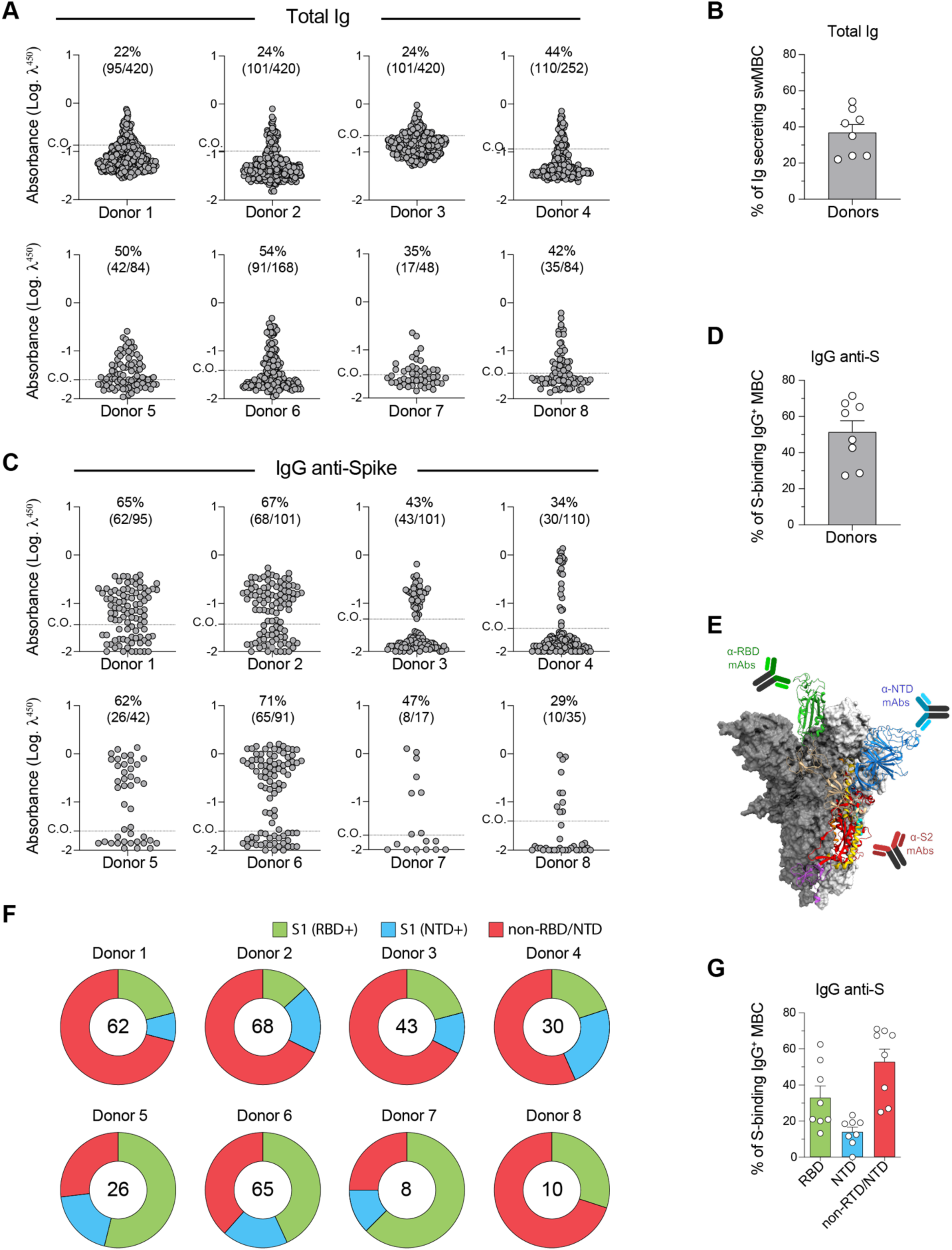
Single-cell cultures (SCCs) for antigen-directed analysis of the Ig repertoire and monoclonal antibody (mAb) isolation. **(A)** Frequencies of antibody-producing cells in SCCs from eight vaccinated donors, ranging from 22% to 54%. **(B)** Pooled data showing frequencies of total Ig-secreting clones across donors. **(C)** Frequencies of mAbs binding to the Wuhan Spike protein, ranging from 29% to 71%. **(D)** Pooled data showing frequencies of Spike-specific clones across donors. **(E)** Schematic representation of the Spike protein regions, including the receptor-binding domain (RBD, green), N-terminal domain (NTD, blue), and S2 region (red). **(F)** Pie charts showing the distribution of mAbs targeting the RBD, NTD, and non-RBD/NTD (undefined) for each donor. **(G)** Pooled frequencies of domain-specific mAbs across donors. Error bars represent SEM.

Among the 592 mAbs produced by responding MBCs, 312 exhibited IgG reactivity against the Wuhan strain Spike protein. Spike-specific clones varied among donors, accounted for 29% to 71% of the Ig-positive wells (**Figure 2C and Supplementary Table 1**). On average, 51% of the clones were identified as S-positive (**Figure 2D**), consistent with our findings from the SCC in **Figure 1H**. Importantly, SCCs secreted on average ∼142 ng/mL of functional Spike-specific IgG, as defined by calibration against a reference anti-Spike IgG human monoclonal antibody (**Supplementary Figure 2C**).

The trimeric SARS-CoV-2 Spike protein consists of three identical subunits, each with distinct functional domains, including the S1 and S2 subunits, as depicted in **Figure 2E**. The S1 subunit contains the receptor-binding domain (RBD), and the N-terminal domain (NTD), while the S2 subunit facilitates the fusion of the virus with the host cell membrane. The RBD is critical for viral entry, as it directly interacts with the ACE2 receptor of human cells, making RBD-specific antibodies highly neutralizing due to their ability to block this interaction. NTD-binding antibodies destabilize the Spike protein trimer, impairing its function and reducing infectivity (21, 22).

To further characterize the obtained mAbs, we analyzed their binding capacity to the RBD and NTD regions by ELISA. Donors exhibited higher serum IgG levels against RBD than NTD (**Supplementary Figure 2D**). Consistently, RBD-specific clones accounted for 33% of cellular responses, compared to 14% NTD-specific clones, aligning with results from previous studies (23). Notably, 53% of clones bound to unknown regions, outnumbered those targeting RBD or NTD (**Figure 2F and G**). We observed a bimodal distribution in the percentage of RBD-specific clones across different donors (**Figure 2G**). For example, donors vaccinated with CoronaVac (CV; donors 1–3) displayed clones that were negative for both RBD and NTD specificity and had higher frequencies of clones recognizing other regions of the Spike antigen, (**Figure 2F**). Donor 4, also vaccinated with CV, had a history of multiple infections with Wuhan and Omicron variants and exhibited broader responses. In contrast, donors vaccinated with AstraZeneca (AZ; donors 5–7), exhibited a higher frequency of RBD-specific clones. Donor 8, also vaccinated with AZ, displayed a higher proportion of clones binding to other regions (**Figure 2F**). When grouped by vaccine type, distinct patterns of clone specificity emerged. Donors primed with CV exhibited significantly fewer RBD-specific clones compared to those primed with AZ, resulting in a higher frequency of clones recognizing other regions of the Spike antigen (**Supplementary Figure 2E**). The observed frequencies of domain-specific clones align with previous reports on SARS-CoV-2-infected cohorts (24), suggesting that the data obtained with SCC method described here accurately represents the underlying antibody repertoire without introducing artifactual biases.

### Comparative cross-reactivity of SCC-derived mAbs to SARS-CoV-2 variants

During the COVID-19 pandemic, identifying mAbs capable of targeting evolutionarily conserved epitopes in the SARS-CoV-2 Spike protein became a major focus due to the virus’s high mutation rate. To evaluate this aspect in our system, we conducted a cross-reactivity study using mAbs derived from SCCs obtained from donors with varying anti-Spike serum IgG levels from different variants (**Figure 3A**). We tested the reactivity of these mAbs against the Spike proteins of four major variants of concern (VOCs): Beta (“B”), Delta (“D”), Gamma (“G”), and Omicron BA.2 (“O”). Across the VOCs, the mAbs exhibited a similar average frequency of immunoreactivity, ranging from 46% to 54%, except for Omicron, which showed a lower average of ∼30%. The mean immunoreactivity frequencies of mAbs for each variant, stratified by donor, are illustrated in **Figure 3B**. While these data provide an overview of variant recognition by mAbs, they do not differentiate between strain-specific and cross-reactive responses.

**Fig. 3:**
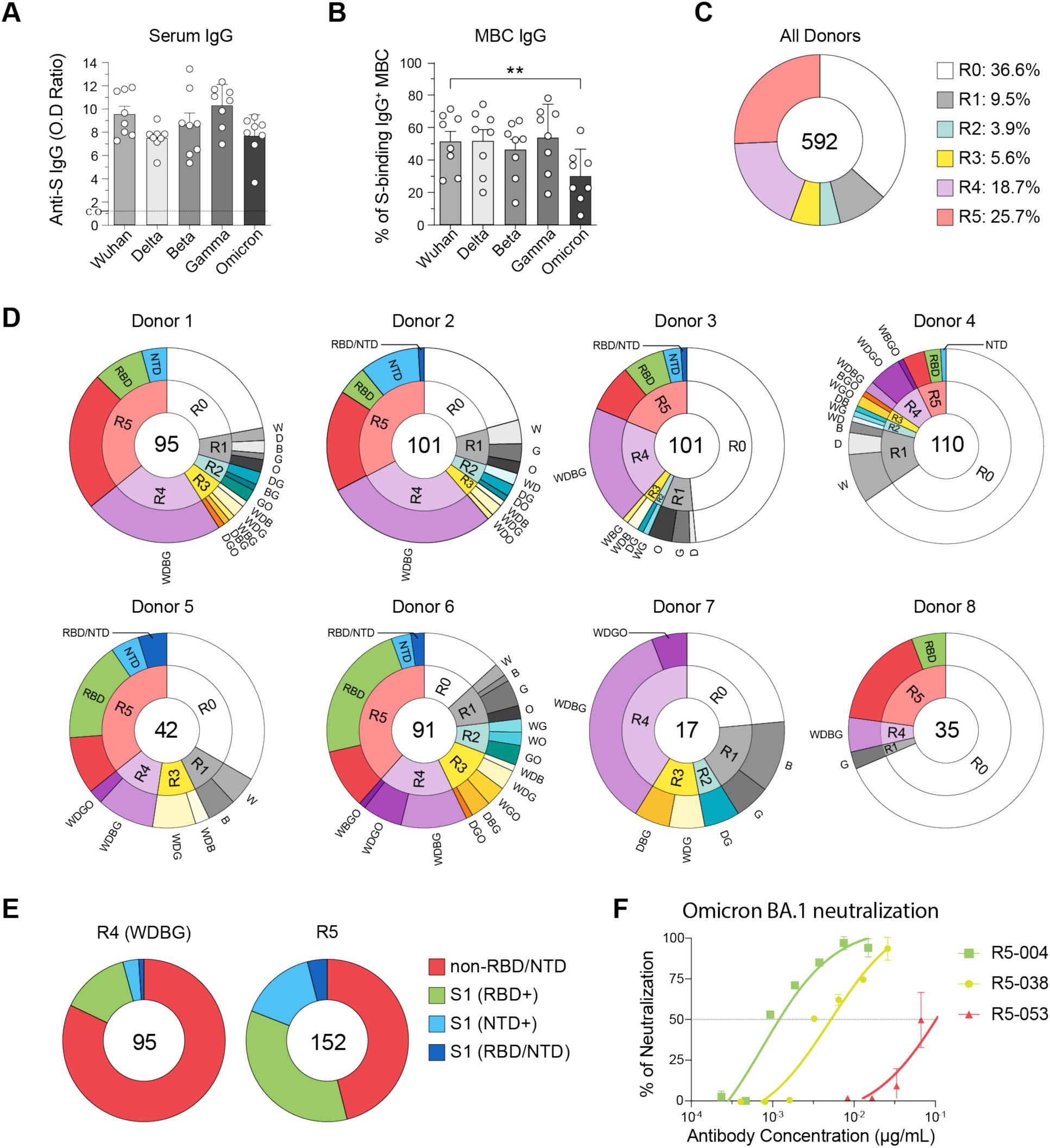
Comparative cross-reactivity analysis of SCC-derived monoclonal antibodies (mAbs) against SARS-CoV-2 spike protein variants. **(A)** Serological analysis showing Spike-specific antibody responses against variants of concern (VOCs): Wuhan, Delta, Beta, Gamma, and Omicron (BA.2). **(B)** Frequency of IgG+ MBCs that recognized each VOC. S+ MBC cells were sorted, cultured and IgG production was assayed by ELISA. VOC reactivity was also measured by ELISA, through plates coated with Spike proteins from each variant. Statistical analyses were performed using one-way ANOVA, and Tukey’s multiple comparison test. Significant difference is indicated by asterisk (**, P < 0.01) **(C)** Monoclonal antibodies isolated from the eight donors, stratified by cross-reactivity levels: R0 (no reactivity), R1 (reactivity to one variant), R2 (two variants), R3 (three variants), R4 (four variants), and R5 (reactivity to all five tested variants). **(D)** Cross-reactivity profiles for each of the eight donors, detailing the distribution of mAbs by cross-reactivity level and highlighting R5 mAbs, which are further stratified by site-specific binding. R5 mAbs are emphasized due to their broader neutralizing potential. **(E)** Pie charts showing the proportions of R4 mAbs (excluding those non-reactive to Omicron, the most genetically divergent VOC) and R5 mAbs, categorized by specificity to the RBD, NTD, or undefined Spike regions (non-RBD/NTD). **(F)** Neutralizing activity of isolated R5 mAbs with site-specific binding to RBD (004 and 038) and putative S2-binding (053).

To further dissect cross-reactivity, we analyzed 592 mAbs isolated from eight donors. Among these, 217 clones showed no binding to any Spike protein variant (R0), and 312 clones were reactive to the Wuhan strain, with 15 clones binding exclusively this strain. A total of 375 clones were categorized into five reactivity groups based on their binding profiles, encompassing 31 distinct and overlapping categories. The R1 group included clones reactive to a single strain, while R2, R3, and R4 groups represented mAbs binding to combinations of two, three, and four strains, respectively. The R5 group comprised mAbs that were reactive to all five tested strains. These reactivity groups are detailed in **Figure 3C and Supplementary Table 1**, with each group assigned labels reflecting the recognized VOCs, for example “WBO” designates clones that bind the Wuhan, Beta, and Omicron variants.

We analyzed the distribution of these categories among donors, with particular emphasis on domain-specific binding among R5 clones, which exhibited the highest degree of cross-reactivity across all tested VOCs (**Figure 3D**). Within the R1 group, we found 15 clones binding exclusively to Wuhan, and 12, 11, 10 and 8 mAbs binding to Gamma, Delta, Omicron, and Beta, respectively. The R1 group, representing strain-specific antibodies, is useful for highly specific immunological assays. R2 and R3 mAbs, which bind two or three strains, are valuable in immunocompetition assays probing inter-variant interactions. R4 antibodies, reactive to all but one variant, help identify signatures of broad binding and structural or genetic factors that impair recognition of particular VOCs, such as Omicron. Comparing R4 and R5 mAbs could reveal key molecular features underlying broad reactivity. Antibodies in the R5 group, which recognize all five variants, likely target highly conserved epitopes, making them particularly valuable for therapeutic applications.

While a distinct distribution of cross-reactive clones was observed across donors (**Figure 3D**), most clones fell into R4 and R5 groups, with 111 R4 clones, 95 of which were specifically classified as R4 (WDBG), and 152 R5 clones (**Supplementary Table 1**). We performed a comparative analysis of mAbs within these two categories, particularly the R4 (WDBG) group, recognizing all variants except Omicron, and the R5 group, which includes antibodies reactive to all five tested strains (**Figure 3E**). Broadly cross-reactive clones are generally expected to target conserved regions of the Spike protein, such as the S2 region, making RBD and NTD less likely targets. This pattern held for R4 (WDBG) antibodies, with 82% of clones binding to the putative S2 region, or other undefined domains (non-RBD/NTD). Surprisingly, 54% of the R5 mAbs bound either RBD (35%), NTD (15%), or both (4%), suggesting that a substantial fraction of these antibodies may possess substantial broadly neutralizing potential.

The cross-reactivity analysis effectively dissects the frequency of strain-specific and cross-reactive mAbs but did not provide insights into the strength or affinity of antibody binding. To address this limitation, we employed a heatmap representation as an alternative approach to visualize the complexity of cross-reactive analysis, providing at least a semi-quantitative assessment of the relative binding strength of each mAb. Binding intensity was evaluated using the ELISA optical density (O.D) ratio, calculated as the antigen-specific O.D. divided by the assay cut-off (defined as the average background signal plus three standard deviations). An O.D. ratio above 1 was considered positive.

**Supplementary Figure 3** illustrates the binding intensity of each R5 mAb isolated to the Spike protein of the tested VOCs, including key neutralizing antibody targets such as the RBD and NTD. Clones were ordered in descending binding intensity to the Wuhan strain and labeled sequentially from R5-001 to R5-152 (**Supplementary Figure 3A**). They were also ranked based on Omicron binding intensity (**Supplementary Figure 3B**). Notably, mAbs exhibiting higher binding intensity for Wuhan Spike were predominantly RBD-specific. This visualization facilitated the identification of clusters of clones with similar binding intensity, enabling the selection of specific candidates tailored to particular study designs. For instance, in the heatmap, we highlight clones chosen for downstream functional analysis, demonstrating the utility of this approach in prioritizing mAbs for further investigation.

### SCC supernatants support neutralization assays and functional mAb candidate selection

To functionally characterize the broadly binding R5 antibodies, we selected three SCC-derived mAbs for pseudovirus neutralization assay against the Omicron variants BA.1 and BA.2: two RBD-binding mAbs (R5-004 and R5-038) and one mAb that does not appear to bind either RBD or NTD (R5-053). The Omicron strain was chosen due to its generally lower intensity of binding observed in prior assays and its high divergence compared to the original Wuhan strain. As shown in **Figure 3F** these mAbs exhibited variable neutralization capacity against the BA.1 variant. The RBD-binding mAb R5-004 demonstrated strong neutralization potential, achieving half-maximal inhibitory concentration (IC50) values in the range of 0.001–0.01 µg/mL. In contrast, RBD-binding mAb R5-038 and the non-RBD/NTD binding clone R5-053 showed moderate to weak neutralizing activity, with IC50 values ranging from 0.01–0.1 µg/mL for the first and a limit of 50% neutralization at the highest concentration used (> 0.1 µg/mL), respectively. Notably, none of the tested mAbs were able to neutralize the BA.2 variant (data not shown), underscoring the increased challenge posed by this more divergent Omicron subvariant. The systematic evaluation of SCC-derived mAbs demonstrates the utility of our platform in mapping cross-reactive and strain-specific antibody responses. These findings lay the groundwork for developing diagnostic and therapeutic tools targeting SARS-CoV-2 and its evolving variants.

### SCC method enables immunoglobulin repertoire analysis and reveals convergent V region rearrangements

Our SCC platform allows to interrogate the clonotypic immunoglobulin repertoire at the transcriptional level. mRNA from expanded SCC-derived MBCs was extracted and subjected to *Ig* heavy and light chain sequencing using V region-specific RT-PCR followed by Sanger sequencing. We sequenced a total of 240 of Ig-secreting MBCs, of which 220 were identified as IgG+ Spike-specific clones. To validate that our culture system did not distort the B cell repertoire, we analyzed the V_H_ gene segment usage and compared the frequencies to a dataset of healthy individuals, and with previous COVID-19 studies (**Figure 4A**). Our findings revealed that gene segments IGHV3-30, IGHV5-51, IGHV3-66, and IGHV1-24 were significantly overrepresented in our cohort, whereas IGHV1-18, IGHV3-23, and IGHV1-69 were underrepresented (**Figure 4A**). Notably, the high frequency of IGHV3-30 aligns with its consistent identification in SARS-CoV-2 antibody repertoire studies (14, 25–28).

**Fig. 4:**
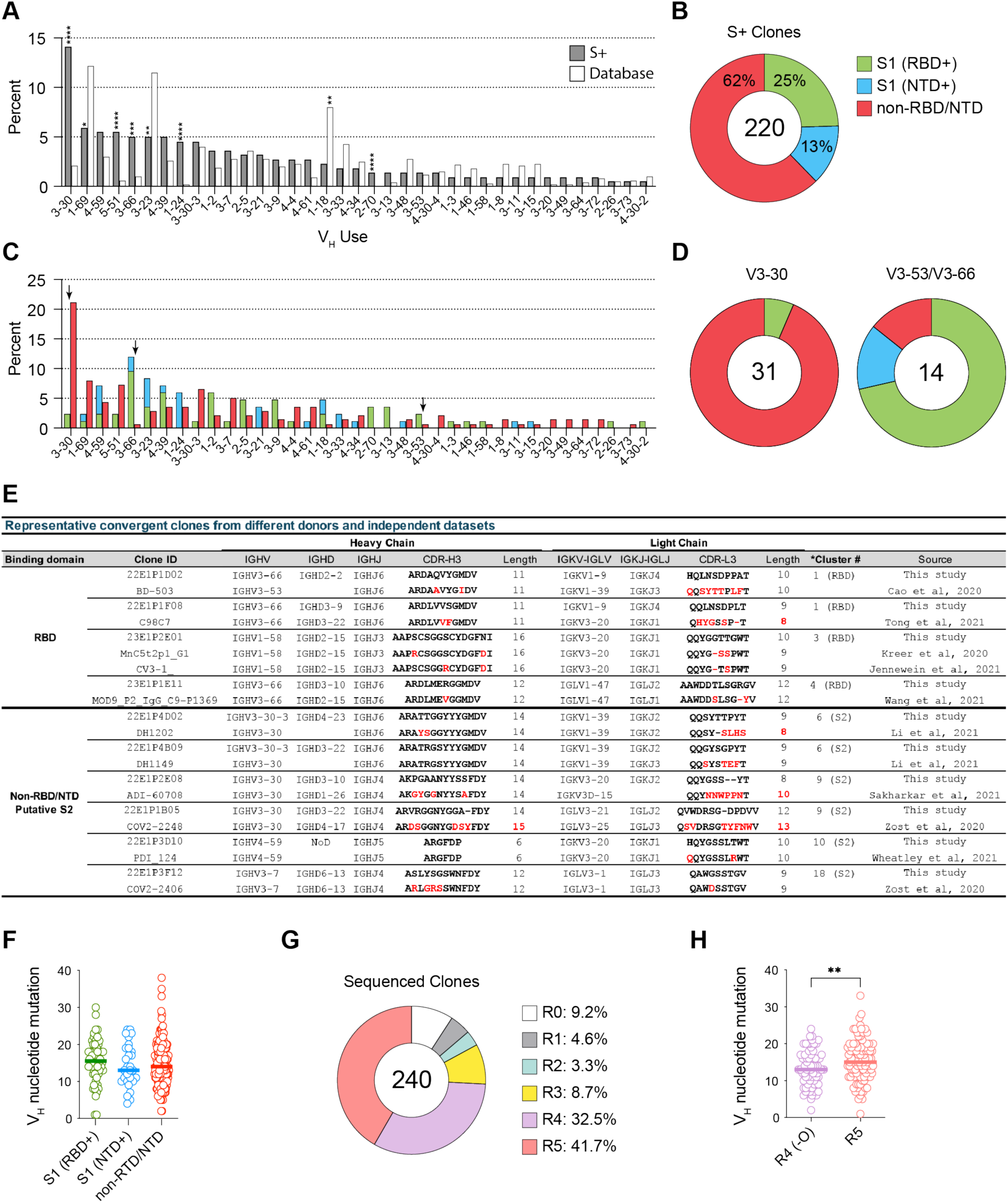
Immunoglobulin repertoire features and convergent V region rearrangements. (**A**) Frequency distribution of IGHV from clones isolated in the current study (gray bars) was compared to the frequency observed in healthy donor dataset (white bars). Statistical significance determined by two-sided binomial test with unequal variance. (**B**) Pie chart showing the percentage of S specific isolated antibodies that bind RBD, NTD or neither. The total number of antibodies tested is indicated at the center of the chart. RBD-/NTD-clones are likely to be S2 binding clones. (**C**) V_H_ usage distribution for clones that bind RBD, NTD or neither. (**D**) Pie charts displaying the percentage of RBD, NTD or non-RBD/NTD binding clones in V3-30 or V3-53/ V3-66 clones. **(E)** Representative convergent antibody clusters. Clones that share 80% of CDRH3 identity as well as VH/VL usage with previously described clones that fall into clusters defined by Wang and colleagues. **(F)** Number of mutations detected in VH genes per clones that bind RBD, NTD or neither. **(G)** Pie chart with the total number of sequenced clones and the percentage distribution for VOC binding, from 0 variants (R0) to all five (R5). **(H)** Comparisons in V_H_ mutations detected per clones that bind four (R4, except Omicron) and five (R5) variants. Statistical analyses were performed using the unpaired two-tailed Student’s t test. Significant differences are denoted by asterisks (*, P < 0.05; **, P < 0.01; ****, P < 0.0001).

Throughout the COVID-19 pandemic, significant efforts were made to identify common features of protective antibody responses against SARS-CoV-2 (26, 29, 30). A seminal study highlighted specific patterns in the immunoglobulin V region, such as V gene usage bias and CDR-H3 composition, associated with binding to RBD, NTD, or S2 regions (27). Here, we compared our data to these previously described patterns to identify convergent antibody responses. Each SCC-derived clone had their mAb-enriched supernatants tested for binding to the full Spike protein and to isolated RBD and NTD domains. This enabled classification of clones into three groups: RBD+ (S1), NTD+ (S1), and non-RBD/NTD (putative S2-or other-region binders). Among 220 S-binding clones, 25% were RBD+, 13% were NTD+, and 62% were RBD-/NTD-(**Figure 4B**).

Efforts to identify potent neutralizing antibodies against SARS-CoV-2 have highlighted the importance of paralogous genes IGHV3-66/V3-53, which are enriched in RBD-binding clones (31–36). In our data, IGHV3-66 was five times more frequent compared to the healthy repertoire (**Figure 4A**). Although only 25% of total clones were RBD+ (**Figure 4B**), 71% of IGHV3-66 clones were RBD+ (**Figure 4C and 4D**).

Conversely, a strong bias toward IGHV3-30 usage has been observed in S2-binding antibodies (27). Consistent with this, most IGHV3-30 clones in our study were classified as RBD-/NTD-(**Figure 4C, 4D**). Light chain V gene usage also exhibited domain-related biases. For instance, IGKV3-20 was associated with S2-targeting antibodies, while IGKV1-9 and IGKV1-33 were prevalent among RBD-binding clones (**Supplementary Figure 4A** and **5A**) (34).

Since our mAbs were tested for RBD and NTD binding via ELISA but not for S2 binding, we cannot experimentally confirm here that the RBD-/NTD-clones specifically target this region. To address this limitation, we performed *in silico* docking studies to assess our presumption. First, we validated the docking approach by submitting two RBD-binding clones for analysis, which confirmed their preferential binding to the RBD region of the Spike protein (**Supplementary Figure 4A**). To investigate whether the putative S2-binding clones indeed targeted the S2 region, we selected four RBD-/NTD-clones for docking analysis, including two encoded by the IGHV3-30 gene segment (22E1P2E08 and 22E1P1B05). The docking results supported their binding preferences for regions within the S2 domain (**Supplementary Figure 4B**), providing further evidence for their classification as S2-binding clones.

We next investigated whether our dataset contained convergent paired IgH/IgL antibody sequences matching those reported in other SARS-CoV-2 studies. Convergent sequences were defined as those sharing the same VH/JH and VL/JL gene usage (allowing minor exceptions for light chains) and exhibiting at least 80% CDRH3 sequence identity. Despite the substantial combinatorial and junctional diversity of the human antibody repertoire, we identified at least 10 clones from our dataset that matched sequences previously reported in independent SARS-CoV-2 studies (14, 27, 31, 37–42). Our analysis revealed four RBD-specific sequences and six putative S2-specific sequences highly similar to clones described in prior datasets (**Figure 4E**), providing further validation of our methodology and supporting the conserved nature of these antibody responses.

The distribution of CDR-H3 amino acid length of mAbs reflected a typical human repertoire (43). Clones classified as RBD⁻/NTD⁻ (putative S2-binding) most frequently exhibited CDR-H3 loops of 14 amino acids, whereas 11 amino acids predominated in RBD⁺/NTD⁺ clones (**Supplementary Figure 5B** and **C**). This pattern aligns with findings from Wang et al., where three major S2-binding clusters had CDR-H3 lengths of 14 amino acids (27). Interestingly, the mean CDR-H3 length for NTD-binding clones was longer averaging 18 amino acids compared to 15 for RBD and RBD-/NTD-clones (**Supplementary Figure 5C**).

Charge distribution within the CDR-H3 loop was similar across groups, with a tendency toward neutrality (**Supplementary Figure 5D** and **E**). No overall differences in amino acid composition were observed between the RBD-/NTD-(S2) and RBD+/NTD+ (S1) groups (**Supplementary Figure 4F**). To delve deeper into potential domain-specific differences, we analyzed clones with biased CDR-H3 lengths, 14 amino acids for the RBD-/NTD-group and 11 amino acids for the RBD+ group, focusing on their amino acid content. Significant differences were observed: RBD-/NTD-(S2) clones with 14 amino acid CDR-H3 loops showed increased use of neutral amino acids such as Tyr, Gly, and Ser, compared to RBD+ clones (**Supplementary Figure 5G and data not shown**). A conserved motif 97[S/G]G[S/N]Y100 was identified in RBD-/NTD-clones such as 22E1P1B05 (**Figure 4E and Supplementary Figure 4B**), consistent with previous studies (27). In contrast, clones with 11-residue CDR-H3 loops showed no clear bias for neutral amino acids in either S1 or S2-targeting groups (**Supplementary Figure 5G**).

The level of somatic hypermutations (SHM) did not seem to differ among clones that target the different domains (**Figure 4F**). The 240 clones were also analyzed for antigenic breadth and classified based on their ability to recognize S protein variants using the cross-reactivity scale, R0-R5 (**Figure 4G**). Importantly, the largest group was R5 (41.7%) followed by R4 (32.5%). The main distinction between R5 and R4 is the ability to recognize Omicron Spike, the most divergent variant tested. Clones recognizing the 5 VOCs, including Omicron (R5), appeared to accumulate more point mutations than those that could not bind Omicron (**Figure 4H**), suggesting a role for SMH and affinity maturation in broadening antibody reactivity. Altogether, these findings establish the SCC platform as a powerful and reliable tool for comprehensive, antigen-specific immunoglobulin repertoire studies, while also highlighting critical conserved features of SARS-CoV-2 convergent antibody responses.

### SCCs enable functional antigen-specific mAb screening, reducing time and resources before scale-up

The SCC methodology effectively identifies antigen-specific and cross-reactive mAbs, streamlining the selection process before large-scale production. As VH/VL sequences can be retrieved from single B cell cloning cultures, this approach enables the subsequent production of large quantities of selected recombinant mAbs for downstream applications, including functional assays, structural studies, and therapeutic development. At the onset of the COVID-19 pandemic, before vaccines were available in Brazil and before the global spread of variants, we isolated several mAbs from patients who recovered from the original Wuhan strain (Cohort 1, **Figure 1**). From this cohort, we selected two broadly reactive RBD-binding clones identified in our 2020 screening assay to evaluate their cross-reactivity and neutralization potential. The Ig genes from these clones were sequenced and expressed in Expi293 cells. Interestingly, these clones expressed convergent V-region rearrangements, including public V3-53 and V3-66 antibodies, consistent with findings that were later published by several groups, (26–28, 31, 36, 37, 44). The selected clones were encoded by VH3-53 (LBL01) and VH3-66 (LBL02), both using VK1-9 for their light chains. The CDR-H3 and CDR-L3 sequences, along with V and J segment usages, are shown in **Figure 5A**.

**Fig. 5:**
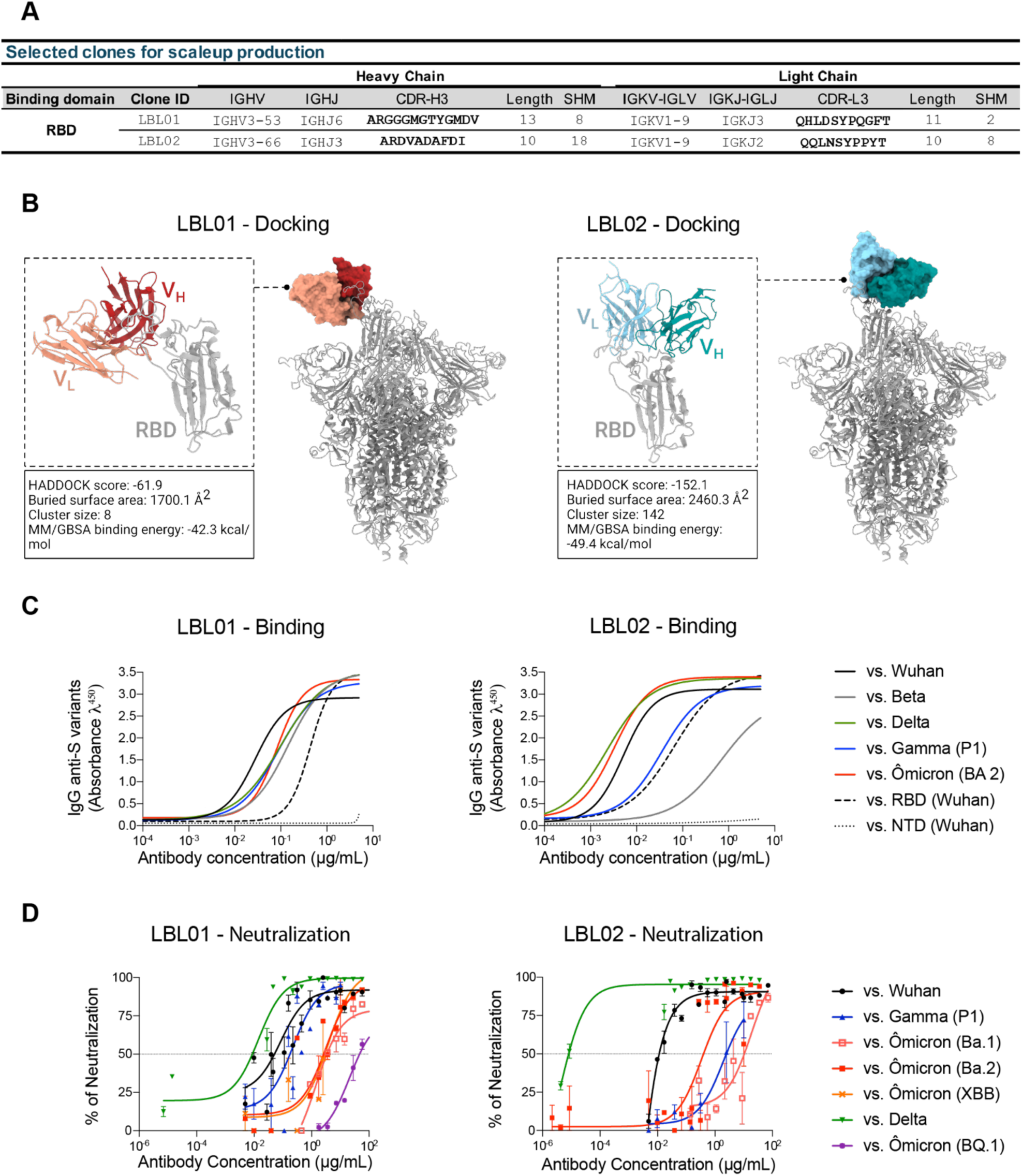
Neutralizing potential of two selected RBD-targeting public clones. **(A)** Heavy and light chain CDR3 sequences of two public clones (LBL01 and LBL02) identified by ELISA as RBD binders. **(B)** Computational docking of each clone, showing the potential RBD binding site for each. **(C)** LBL01 and LBL02 antibodies were produced and tested in ELISA against five VOCs, as well as RBD and NTD domains. **(D)** Pseudovirus neutralization assay for both clones, confirming that the broader specificity of LBL02 is also functionally reflected in its stronger neutralization capability

Computational docking analysis corroborated the binding of both clones to the RBD *in silico* (**Figure 5B**). Further blind docking analyses against multiple VOCs revealed limited cross-reactivity for LBL01, whereas LBL02 exhibited the potential to a broader reactivity as distinct docking runs converging on a similar binding site (**Supplementary Figure 6**). Accordingly, molecular dynamics simulations of the refined interaction poses followed by binding free energy (**ΔG_bind_**) calculations, revealed a stronger interaction of LBL02 with the RBD (−49.4 kcal/mol) compared to LBL01 (−42.3 kcal/mol), suggesting a more stable binding interface that may confer greater resistance to viral escape (**Figure 5B**; **Supplementary Figure 6**).

The two mAbs were then produced at scale from Expi293 cultures, purified, and tested for binding to S proteins from Wuhan, Delta, Beta, Gamma, and Omicron (BA.2), as well as to the isolated RBD and NTD domains (**Figure 5D**). Both antibodies were confirmed to target the RBD. Titration experiments demonstrated that LBL01 exhibited relatively weak binding to VOCs S proteins compared to LBL02, which displayed strong apparent affinity across all tested variants. Pseudovirus neutralization assays were performed against Wuhan and VOCs, including Delta, Beta, Gamma, and Omicron (BA.2, BA.1, and XBB). Consistent with binding and *in silico* analyses, LBL01 showed weaker neutralization against Delta, BA.1, and Wuhan strains but retained limited activity against the Omicron subvariants BQ.1 and XBB. In contrast, LBL02 strongly neutralized Wuhan and Delta but showed sharply reduced activity against Omicron subvariants (**Figure 5E**).

These findings suggest that some early-pandemic immune responses may have conferred cross-protection against emerging variants, emphasizing the importance of rapidly identifying broadly neutralizing antibodies for future pandemic preparedness.

Altogether, these findings demonstrate that the SCC platform enables efficient selection of clones with desired specificity and functional neutralizing potential prior to scaling up production. This method reduces the time and resources typically required for traditional cloning and expression workflows, providing a streamlined and cost-effective approach for monoclonal antibody discovery.

## Discussion

Memory B cells are essential components of the adaptive immunity, enabling rapid and robust responses upon re-exposure to pathogens. While serum antibody titers provide a snapshot of circulating immunity, MBCs represent the capacity for secondary responses upon antigen re-exposure, providing a dynamic perspective on immune memory. By isolating and characterizing these cells, researchers can explore the mechanisms underlying long-term immunity, yielding valuable insights for vaccine and immunotherapy development. Such studies are particularly critical for emerging infectious diseases, where timely and precise immune responses can be pivotal in controlling outbreaks. MBC-derived mAbs have revolutionized treatment strategies for diseases like COVID-19. These studies not only deepen our understanding of immunity but also provide practical tools for combating infectious and autoimmune diseases (37, 45, 46).

We previously developed a method for studying the mouse B cell repertoire, combining high-efficiency B cell cultures supported by feeder cells with single-cell RT-PCR amplification of Ig variable genes and sequencing (10). This approach provides access to the entire naïve and memory B cell repertoire, simultaneously enabling antigen specific analysis and VH/VL repertoire usage. Here, we adapted this strategy to analyze the human MBC repertoire and established a streamlined pipeline for isolating specific human monoclonal antibodies. Our goal was to democratize access to comprehensive humoral immune profiling, enabling a wide range of laboratories to address local infectious disease challenges with innovative solutions to support both immunological and translational studies.

It is noteworthy that all the feeder cells used in our study were derived from mice, yet they effectively support the growth and differentiation of human B cells in culture. This cross-species compatibility suggests that key survival and activation signals provided by these feeder cells may be shared across species. Among the tested feeder cell lines, NB21 cells, engineered to secrete IL-21, were the most effective in supporting MBC survival and promoting immunoglobulin secretion. IL-21 boosts immunoglobulin production, particularly IgG, which is crucial for functional antibody repertoire analyses. While the prevailing protocols for culturing human MBCs with feeder cells rely on complex cytokine cocktails (12, 13), our culture system employs a simplified yet effective strategy inspired by Pinna et al. (2009) (8). These authors optimized human MBC proliferation and Ig secretion using R848 or CpG combined with high-dose IL-2 (8). Given R848’s cost-effectiveness and dual activation of TLR7 and TLR8, here we chose R848 as a polyclonal stimulus in our culture system. Therefore, by combining NB21 cells, R848 polyclonal stimulation, and IL-2 supplementation, we successfully establish here a robust and reproducible platform for both quantifying the frequency of antigen-specific human MBCs clones and characterizing their VDJ/VJ repertoire.

Our data show that the methodology is sensitive enough to detect and characterize antigen-specific MBC responses in both SARS-CoV-2-exposed donors and convalescent patients. The results underscore the sensitivity and reliability of the proposed platform for studying and comparing MBCs responses across individuals and experimental groups using a standardized feeder cell system, thereby assuring reproducibility. The ability to capture low-frequency antigen-specific MBCs is relevant for understanding immune memory, given the vast antigenic repertoire that MBCs must encode over a lifetime.

Traditional approaches often struggle to reveal these rare populations due to their low abundance. Here we show that the antigen-specific repertoire can be probed using LDA cultures. Our methodology demonstrated sufficient sensitivity to distinguish SARS-CoV-2-exposed donors from convalescent patients sampled in 2020. Interestingly, we observed a correlation between Spike-specific MBC frequencies and serum anti-Spike IgG levels. Similar correlations have been reported in previous studies of patients exposed to antigens through immunization or infection (5). This correlation supports the use of MBC profiling as a complementary tool for immunoepidemiological surveillance of pathogen exposure and immune memory.

To facilitate the isolation of antigen-specific MBCs for mAb production, we employed a baiting strategy using Wuhan strain Spike protein conjugated to two different fluorescent markers (**Figure 1 C**). We showed that SCCs of bait-sorted cells yielded total percentages of responding B cells comparable to those obtained through LDA of total MBCs, while significantly enhancing the isolation of antigen-specific mAbs. These results demonstrate that bait-selected B cells maintain viability and are not prone to cell death potentially induced by B-cell receptor cross-linking. Notably, the baiting strategy increased the yield of antigen-specific mAbs by 30-to 50-fold compared to cultures of MBCs isolated without antigen baiting.

A key aspect of our SCC platform is the ability to rapidly generate monoclonal antibodies to interrogate multiple specificities within a single antibody sample. Despite the limited supernatant volume recovered from SCCs, we demonstrated the feasibility of performing pseudovirus neutralization and multiple ELISA assays to identify antigen-specific responses and prioritize the most promising mAbs to sequencing. This is particularly beneficial in scenarios where logistical or financial constraints prevent the sequencing of a large number of mAbs. Compared to traditional hybridoma and recombinant mAbs techniques, SCCs offer a crucial advantage by streamlining the identification of antigen-specific clones, eliminating labor-intensive and financially expensive steps such as massive gene sequencing and recombinant mAb expression. These findings highlight the utility of our SCC-based approach as a time-efficient and cost-effective solution for both antibody repertoire analysis and monoclonal antibody discovery.

The Spike protein is the most immunodominant antigen of SARS-CoV-2. Most antibodies targeting Spike recognize the S1 subunit with anti-RBD antibodies accounting for nearly 90% of the neutralizing antibodies found in convalescent plasma from COVID-19 patients (47). The RBD is well established as a critical target for neutralizing antibodies due to its role in binding ACE2 receptor, a key step in viral entry (48). However, the NTD of the S1 subunit also contains an antigenic “supersite” that serves as a major target for antibody responses (49). Notably, as SARS-CoV-2 has evolved, the S1-NTD has accumulated numerous mutations, particularly deletions, which can alter antigenicity or eliminate key epitopes. These alterations enhance the virus’s ability to evade immune recognition, further complicating the maintenance of effective immunity through vaccination or natural infection (50–53). The RBD, especially in variants of concern (VOCs) such as Delta and Omicron, is under intense mutational pressure, posing a significant challenge for vaccine-induced and natural immunity. Antibodies binding to evolutionary conserved epitope within RBD that can recognize all VOCs hold great potential for cross-protective immunity. Similarly, the NTD plays a multifaceted role in viral attachment and entry, but is also prone to mutations and deletions that alter antigenicity and enable immune evasion (53). These structural changes in both RBD and NTD contribute to the virus’s ability to escape recognition, complicating vaccine efficacy and the design of therapeutic antibodies.

In the present work, we isolated 592 mAbs, 312 of which recognized the Wuhan strain Spike protein. Given the well-known immunodominance of the RBD and NTD of the Spike protein (47, 48, 53), we classified mAb specificities within our collection accordingly. Interestingly, the majority of isolated mAbs in this study were non-RBD/non-NTD specific, suggesting that these antibodies may target other S1 conformational epitopes or conserved regions on the S2 subunit, areas not directly tested in our assay. The S2 region, which mediates viral fusion, is less prone to mutations than S1, making it an attractive target for broad-spectrum neutralizing antibodies. The high proportion of non-RBD/non-NTD antibodies observed in our study, is consistent with the predominance of non-RBD/non-NTD antibodies reported in the literature (24). These findings emphasize the importance of expanding the scope of epitope targeting to include non-RBD/non-NTD conserved regions for the development of universal vaccines and therapeutics against SARS-CoV-2 and its variants.

The NTD of the SARS-CoV-2 Spike protein is susceptible to frequent mutations and deletions. These structural changes, commonly observed in several VOCs including Delta and Omicron, significantly contribute to immune escape, thereby impacting vaccine efficacy and natural immunity. Here, we utilized the NTD protein from the Wuhan strain in the ELISA assays. As expected, our analysis revealed that, on average, the NTD region was less frequently recognized by the isolated monoclonal antibodies compared to the RBD and other unidentified regions, in agreement with previous studies. Interestingly, a bimodal distribution of RBD-specific clone percentages among donors became evident when grouped by vaccine type. Most CoronaVac (CV) vaccine recipients displayed a higher proportion of clones recognizing non-RBD/non-NTD regions, whereas 3 out of 4 AstraZeneca (AZ) vaccine recipients exhibited a greater frequency of RBD-specific clones. This variation likely reflects differences in vaccination regimens, prior exposure to diverse viral variants, or underlying genetic heterogeneity among donors.

The cross-reactivity analysis performed here, across Spike proteins, revealed distinct patterns of strain-specific and broadly reactive antibodies, offering valuable insights into the immunological landscape of SARS-CoV-2 and its variants (50–53). Notably, we successfully identified several R5 mAbs that bind to all tested variants. Throughout the COVID-19 pandemic, a key priority has been the identification of mAbs that can target conserved regions of the SARS-CoV-2 Spike protein due to their therapeutic potential. R5 antibodies, which recognize all tested variants, likely target highly conserved epitopes, making them particularly promising candidates for broad-spectrum antiviral strategies.

These conserved binding sites suggest a greater potential for neutralization breadth, a critical attribute given virus’s high mutational rate and the emergence of variants with enhanced transmissibility or immune escape. Antibodies targeting such sites could be instrumental for developing treatments and vaccines that are effective across multiple strains, enhancing the prospects for broader and more durable immunity, offering a critical advantage in controlling outbreaks and mitigating the risk posed by future variants.

Comparative analyses between R4 and R5 clones may further illuminate the molecular determinants of broad reactivity, contributing to the design of next-generation immunotherapies. In parallel, strain-specific antibodies, such as those in the R1 group, also hold significant value as precise tools for detecting individual variants, making them useful for variant-specific diagnostic assays.

Broadly reactive clones were isolated in the present study through SCC supernatant screening, including R5-004 and R5-053. As a proof of concept to demonstrate the method’s capability to use supernatants containing mAbs to identify clones with potential for virus neutralization, we cloned and expressed two public RBD-specificantibodies, LBL01 and LBL02. Functional assays performed with the recombinant antibodies were in full agreement with those obtained with their respective SCC-derived supernatants. These results validate the platform’s utility for the characterization and isolation of antibodies with therapeutic and diagnostic applications.

The high cloning efficiency of our SCC system with NB21 feeder cells enabled unbiased profiling of memory B cells (MBCs) repertoire, revealing convergent rearrangements in immunoglobulin variable (V) regions that align with findings from previous SARS-CoV-2 studies (26, 27, 54). Key aspects include the representation of VH gene segments, particularly IGHV3-30 and IGHV3-53/IGHV3-66, and their association with distinct Spike regions. Importantly, VH/VL analysis of SCC-derived clones exhibited somatic hypermutations correlating with cross-reactivity, particularly for challenging variants like Omicron. Consistent with earlier studies, IGHV3-53/IGHV3-66 was strongly linked to RBD-binding antibodies, while IGHV3-30 was primarily associated with putative S2-targeting clones (27). Computational docking confirmed these classifications, providing evidence for conserved binding motifs and structural preferences. The paralogous genes IGHV3-53 and IGHV3-66 are frequently described as encoding public clones across diverse populations (36, 55). Interestingly, while most global studies report a higher prevalence of IGHV3-53 compared to IGHV3-66, our dataset showed a fourfold higher frequency of IGHV3-66 among RBD-binding antibodies. This distinctive finding might reflect a population-specific features of the Brazilian cohort, as a similar preference for IGHV3-66 was observed in another study involving Brazilian blood donors (25).

Limitations of this study include the use of Wuhan strain-based reagents, which may underestimate responses to more recent variants. Additionally, the classification of mAbs as non-RBD/non-NTD was based on lack of reactivity rather than confirmed S2 binding, underscoring the need for further site-specific targeting. While *in silico* analyses support these assignments, structural validation would strengthen the conclusions. The SCC methodology, although effective for identifying antigen-specific and cross-reactive mAbs, yields limited antibody quantities, sufficient only for initial screening. However, because VH/VL sequences can be retrieved from SCCs, this limitation can be addressed through recombinant expression of selected clones. Finally, the small number of patients studied here limits our ability to draw conclusions about the clonal distribution of memory B cell repertoires and their persistence following vaccination. With expanded cohorts, however, this approach may prove valuable as a novel tool for immunoepidemiological surveillance.

In summary, this study presents a reliable methodology for isolating clinically relevant monoclonal antibodies. By enabling functional screening of antigen-specific mAbs directly from culture supernatants, our SCC platform provides insights into binding properties, site specificity, cross-reactivity, and neutralization potential. It simplifies mAb discovery by reducing the need for large-scale sequencing and early recombinant antibody production, thereby lowering costs and labor. This strategy improves workflow efficiency by prioritizing clones for downstream applications based on functional data. Its affordability and adaptability make it valuable for laboratories worldwide, particularly in low- and middle-income regions, by equipping researchers with practical tools for immunological analysis and therapeutic development. Importantly, the platform opens new avenues for immune-epidemiological investigations beyond serology, allowing studies that link the quality and magnitude of memory B cell responses with protection, serum titers, and clonal repertoire composition.

## Materials and Methods

### Ethic Statement

This study was approved by the National Ethics Committee (CONEP, Brazil; CAAE: 30161620.0.0000.5257) and the local ethics committee of Fundação Oswaldo Cruz (CAAE: 3048.7120.2.0000.0008). Blood collection from the volunteers in this study was carried out at the Diagnostic Center of Núcleo de Enfrentamento e Estudos de Doenças Infecciosas Emergentes e Reemergentes (NEEDIER) located at the Universidade Federal do Rio de Janeiro and at Fundação Oswaldo Cruz, in adherence to the approved protocols.

### Study Participants

For the first part of the study, involving Limiting Dilution Assay (LDA) culture experiments, we enrolled fourteen patients from Brazil, collecting whole blood samples at NEEDIER to perform Limiting Dilution Assay experiments. For single cell culture experiments for monoclonal antibody isolation, we enrolled eight vaccinated and convalescent patients as described in **Supplementary Figure 2 A**. These patients were selected through Fundação Oswaldo Cruz, with whole blood samples collected for further analysis. Participants were categorized into three cohorts. The first cohort consisted of healthcare workers with consecutive negative PCR results (negative exposure group, *n* = 5). The second cohort included non-vaccinated convalescent individuals, sampled 20 to 180 days after their first negative PCR result (*n* = 9). Samples from these two cohorts were collected in 2020, before COVID-19 vaccines became available. The third cohort comprised individuals with multiple vaccinations and convalescent from prior SARS-CoV-2 infection (*n* = 8). Additionally, as a control we included 23 pre-COVID plasma samples collected at the State Hematology Institute Hemorio followed a protocol approved by the local ethics committee (CEP Hemorio; approval #4008095).

### Blood sample processing

All blood samples were collected in tubes with heparin as an anticoagulant and sent to the Laboratory of Lymphocyte Biology (LBL) for subsequent PBMC isolation. To obtain the desired cell population, we followed the purification protocol using Ficoll Histopaque®-1077 (SIGMA-Aldrich cat.#10771). After allowing the sample to settle for 40 minutes at RT, the tubes containing the sample were centrifuged at 1500 rpm for 5 minutes at 24°C. Next, the plasma was removed and frozen, and the blood was carefully transferred to a 15 mL falcon tube containing 5 mL of Ficoll at a 2:1 dilution. This tube was centrifuged again, this time for 30 minutes at 400g, 24°C, without brake or acceleration. The PBMC layer was then collected, washed with 1x PBS, and centrifuged at 1300 rpm for 5 minutes at 24°C. After this step, the pellet was resuspended in 2 mL of ACK Lysing Buffer® (GIBCO, #A10492-01) to lyse red blood cells, keeping the sample in the refrigerator for 5 minutes. After a wash step as described previously the cells was resuspended in FACS Buffer containing the labeled antibodies for staining.

### ELISA for detection of serum immunoglobulins

For quantitative analysis of anti-SARS-CoV-2 spike protein IgG antibodies, we performed the S-UFRJ test, as described previously (56). Briefly, high binding ELISA plates were coated with 50 μL of SARS-CoV-2 spike protein (4 μg/mL in PBS) and incubated overnight. The coating solution was removed and 100 μL of PBS 1% BSA (blocking solution) was added and the plate was incubated at room temperature (RT) for 1-2 hours. The blocking solution was removed and 50 μL of diluted 1:40 (PBS 1% BSA) patient sera were added, subsequently, the sera were serially three-fold diluted in the plate, which was incubated at RT for 2 hours. Then, the plate was washed with 150 μL of PBS (5x) and 50 μL of 1:10000 goat anti-human IgG (Fc)-horseradish peroxidase antibody (Sourthen Biotech, Birmingham, USA) were added, and the plate was incubated for 1.5 hours at RT. The plate was washed again with 150 μL of PBS (5x) and then treated with TMB (3,3’,5,5-tetramethylbenzidine) (Scienco, Brazil) until the reaction was stopped with 50 μL of HCl 1N. The optical density (OD) was read at 450 nm with 655 nm background compensation in a microplate reader (Bio-Rad Laboratories, Inc, California, USA). Relative antibody levels were determined based on a cut-off (C.O) value calculated as the mean absorbance of the negative controls plus one standard deviation on the same ELISA plate. Samples with absorbance values below the C.O were classified as negative, while those with values at least two times above the C.O were considered positive.

### Feeder cell lines preparation

The feeder cells are cultured in T175 flasks (Falcon, Ref #353112) until they reach confluence, using RPMI (Gibco, #11875-093) supplemented with 1% pyruvate (Gibco, #11360-070), 1% glutamine (Gibco, #25030-081), 1% penicillin-streptomycin (Gibco, #15140-122), 1% HEPES (Gibco, #15630-080), 50 nM 2-Mercaptoethanol (Sigma, #M3148), and 10% fetal bovine serum (FBS, Gibco, #10437-028). The cells are passaged every three days. Upon reaching confluence, the cells are detached by treating them with trypsin (Gibco, #15400-054) for 5 minutes at 37°C and 5% CO₂. The culture medium is collected, and the cells are washed with PBS. After centrifugation at 300g for 5 minutes, the pellet is resuspended in 10 ml of RPMI. The cells are then counted using a Neubauer chamber. For plating, 3000 NB21 feeder cells are seeded per well in 100 µl of RPMI with the aforementioned supplements and allowed to grow overnight. The following day, the cells are irradiated with 20 Gy to halt the cell cycle. This irradiation is performed at the Laboratório de Instrumentação Nuclear (LIN), Centro de Tecnologia da UFRJ.

### Cell sorting

The cells were stained in supplemented RPMI using CD19 – BV421, CD38 – Pecy5, CD27 – PE, IgD – Pecy7, DUMP(CD3/CD14/CD16) – APCcy7 and Spike-protein labeled with Alexa 488 and Alexa 647 to access the swMBC population (CD19^+^CD38^-^CD27^+^IgD^-^) and antigen-specific swMBC populations (CD19^+^CD38^-^CD27^+^IgD^-^Spike^APC+FITC+^). Cells were sorted using MoFlo (Dako cytomation) in 5ml tubes with supplemented RPMI.

### B cell cloning cultures

Sorted cells were counted and plated using different densities of cell in LDA or 1 cell per well in SCC. In LDA, since the number of B cells were variable among the patients, we plated in the first row 500 to 2000 cells in eleven columns of a 96 well culture plate flat bottom descending in a 3 fold dilution in the first 4 rows of the plate, the for last rows were plated to tending to zero so it can be possible to calculate Poisson distribution.

We plate the 100µl with the determined number of MBC (LDA or SCC) over a monolayer of NB21 cells and supplemented with the polyclonal stimuli of IL-2 (10nM) and R848 (2µg/ml). In this comparison, R848 (Resiquimod) (Invivogen, #tlrl-r848) and IL-2 (Immunotools, #11340025) were used as polyclonal stimuli, based on previous reports demonstrating their effectiveness in human B cell cultures (8). The cells were left to grow for 5 days, then we collect 25µl of the supernatant to perform the ELISA for detect total immunoglobulin. In the day 6, we collect another 15µl to perform the antigen-specific ELISA. We stop the culture in day 7, removing and storage the supernatant with 0.02% Azide in another 96 well plate. And the B cells were harvested with 200µl of cold PBS 1x and transferred to a PCR plate with 10 µl of TLC + 5% 2-Mercaptoethano for Immunoglobulin gene sequencing.

### ELISA for total immunoglobulins in LDA

High binding ELISA 96-well microplates (Corning) were coated with 50 µL per well using 1:1000 dilution of Ig unlabeled (Southern Biotech #) in PBS 1x for 4°C overnight. In the next day, plates were blocked with 200µl of blocking buffer (PBS 1x with 1% of BSA) for 1h at RT. The blocking buffer was removed, and supernatant was plated diluted two times using blocking buffer in a total volume of 50µl for 2h at RT. The plates were washed five times with 200µl per well of PBS 1x. We used an Ig conjugated with HRP as a secondary antibody using a concentration of 1:4000 dilution in blocking buffer in 50µl per well. The plates were washed as described above and developed with 50 µL of TMB (Scienco, Brazil). The reaction was stopped with 50 µL of 1 N HCl, and the optical density (O.D.) was read at 450 nm with 655 nm background compensation in a microplate reader (BioRad). Positive antibody levels were determined based on a cut-off (C.O) value calculated as the mean absorbance of the negative controls plus three standard deviation on the same ELISA plate. Supernatant with absorbance values below the C.O were classified as negative, while those with values above the C.O were considered positive.

### Antigen-specific ELISA

Half-area High binding ELISA 96-well microplates (Corning) were coated with 30 µL per well using 4µg/ml of the Spike-antigen (Wuhan, Delta, Beta, Gamma and Omicron BA2), produced as previously described (56) or 1µg/ml of available RBD or 1µg/ml of NTD in PBS 1x incubated overnight 4°C. In the next day, plates were blocked with 150µl of blocking buffer (PBS 1x with 1% of BSA) for 1h at RT. The blocking buffer was removed, and supernatants were plated diluted 1:2 using blocking buffer in 30µl volume for 2h at RT. For experiments designed to quantify anti-Spike IgG secreted in SCC supernatants, a standard curve was prepared using serial 1:3 dilutions of an anti-Spike (Wuhan) human IgG monoclonal antibody (CZmAb) of known concentration. The plates were washed five times with PBS 1x. We used an IgG conjugated with HRP as a secondary antibody using a concentration of 1:4000 dilution in blocking buffer in 30µl per well. The plates were washed as described above and developed with 30 µL of TMB (3,3’,5,5-tetramethylbenzidine) (Scienco, Brazil). The reaction was stopped with 30 µL of 1 N HCl, and the optical density (O.D.) was read at 450 nm with 655 nm background compensation in a microplate reader (BioRad). To maximize our resources, we performed the antigen-specific ELISA assays using half-area ELISA plates, which allowed us to save the supernatant and conduct more assays. For the culture supernatants, Positive antibody levels were determined based on a cut-off (C.O) value calculated as the mean absorbance of the negative controls (NB21 culture supernatants without B cells) plus three standard deviation on the same ELISA plate. Samples with absorbance values below the C.O were classified as negative, while those with values above the C.O were considered positive.

### Production of SARS-CoV-2 variant pseudo-typed viruses and pseudovirus neutralization assays

SARS-CV-2 encoding plasmids: pcDNA3.1-SARS-CoV-2-S (Wuhan) was a gift from Thomas Gallagher (57); pcDNA3.3_CoV2_P1 (Gamma), B.1.617.2 (Delta), XBB, and BQ.1 were gifts from David Nemazee (58); pcDNA3.1-CoV2-Omicron BA.1 spike was a gift from Ben Murrell (59). Plasmids encoding the WT (Wuhan) SARS-CoV-2 spike, VOCs (Delta and P.1 Gamma) spikes and VOC Omicron sub-lineage spikes (Ba.1; Ba.2; XBB; BQ.1) were transfected with Lipofectamin 3000 (Thermo) in Hek-293T cells and 24 hours later infected with a VSV-G pseudo-typed ΔG-luciferase (G^∗^ΔG-luciferase) with a multiplicity of infection of 5 for 2 h before washing the cells with 1×PBS. Transfected/infected cells were incubated with fresh DMEM with 5% FCS. The next day, transfection/infection supernatants were collected and clarified by centrifugation at 300g for 10 min before filtration through a 0.45μm filter. SARS-CoV-2 pseudotyped ΔG-luciferase viruses were titrated in Vero E6 cells and aliquots stored at −80°C.

Neutralization assays were performed by incubating pseudoviruses with serial dilutions of sera or mAbs present in the supernatants from SCC monoclonal antibodies and scored by the reduction in luciferase gene expression. In brief, Vero E6 cells were seeded in a white 96-well plate at a concentration of 2×10^4^ cells per well. Pseudoviruses were incubated the next day with serial dilutions of the test samples in triplicate for 30 min at 37°C. The mixture was added to cultured cells and incubated for an additional 24 h. The luminescence was measured by Luciferase Assay System (Promega). IC_50_ was defined as the dilution at which the relative light units were reduced by 50% compared with the virus control wells (virus + cells) after subtraction of the background in the control groups with cells only. The IC_50_ values were calculated using nonlinear regression in GraphPad Prism.

### Immunoglobulin sequencing from single cell cultures

To obtain single cell lysates, at day 6-7 post SCC, MBC were harvested from co-culture 96 well plates, centrifuged and suspended in 10 µL of TCL buffer (Qiagen) containing 1% 2-mercaptoethanol (60). Cell lysate plates were immediately frozen and kept at −80°C for mRNA preservation. RNAClean XP (Beckman Coulter) was used to purify mRNA following the manufacturer’s instructions. cDNA was synthesized with 25 pmol Oligo-dT primer and 50 U of Superscript III reverse transcriptase (Invitrogen) in the presence of 4 U RNAsin (Promega). Reverse transcription was carried out at 42°C for 10 min, 25°C for 10 min, 50°C for 60 min and 94°C for 5 min (7). cDNA was stored at −20°C and variable region from heavy and light chains were amplified using 4µL cDNA and primers presented in **Supplementary Table II** (61). PCR reaction was performed with 1.25 U/ reaction of GoTaq Flexi DNA Polymerase (Promega) in the presence of 25 pmol primers, 0.2 mM dNTPs and 2 mM MgCl_2,_ as suggested by the manufacturer. Cycle conditions were adapted from a previously published study (61), consisting of: 2 min at 94°C, 4 cycles of 30 s at 94°C, 30s at 50°C and 2 min at 72°C, followed by 8 cycles of 30 s at 94°C, 30 s at 55°C and 2 min at 72°C and 28 cycles of 30 s 94°C, 30 s 60°C and 2 min at 72°C. Final extension was performed at 72°C for 10 min. Heavy and light chain PCR products were purified with ExoSAP-IT (Invitrogen) according to thes manufacturer’s instructions and sequenced with 3.2 pmol of the reverse primer. Sanger Sequencing were performed by the PSEQDNA-UFRJ facility, Biophysics Institute, Federal University of Rio de Janeiro.

### Analysis of immunoglobulin sequences

Obtained sequences were converted to fasta file and submitted to gene annotation using IMGT/HighV-QUEST tool (62), in some cases Vbase 2 (63) and IgBLAST were used for confirmation. VH, DH, JH, VK, VL, JK and JL gene segments were annotated, and the number of nucleotide mismatch mutations were calculated. The human V genes usage of the antibodies from this study were analyzed as described by Agudelo et al (25). Briefly, the frequency distributions of human V genes in anti-SARS-CoV-2 antibodies compared to 131,284,220 IgH and IgL sequences generated by Soto et al. (64) and downloaded from cAb-Rep (65), a database of human shared BCR clonotypes available at https://cab-rep.c2b2.columbia.edu/. The two-tailed t test with unequal variance was used to compare the frequency distributions R of human V genes of anti-SARS-CoV-2 antibodies from this study to Sequence Read Archive SRP01097039. CDR-H3 analyses were performed as previously described (66). CDRH3 logograms were plotted using the online tool https://weblogo.berkeley.edu/logo.cgi (67). Average hydrophobicity of CDR-H3 was calculated as previously described (68).

### Antibody expression and purification

The VH and VL antibody sequences of LBL01 and LBL02 were ordered as gBlocks (Integrated DNA Technologies) and cloned by Gibson assembly (New England Biolabs) into a customized pcDNA 3.4 vector containing a human IgG1 Fc region. VH and VL plasmids were mixed at 1:2 ratio and were transfected into Expi293F cells (Thermo Fisher Scientific), which were cultured at 37 °C and 8% CO2 for 6 days, then harvested, centrifuged 4500 x g for 20 min. Antibodies were isolated from filtered (0.22µm) supernatants using Protein G Plus Agarose (Pierce Thermo Fisher Scientific) affinity chromatography, washed with 20 column volumes of PBS, eluted with 150 mM glycine-HCl pH 2.7, and neutralized with 1 M Tris-HCl pH 8.0. The antibodies were buffer exchanged into PBS and concentrated using Pierce™ Protein Concentrators PES 30.000 MWCO centrifugal spin columns (Thermo Fisher Scientific).

### *In silico* analysis of antibody binding

Antibodies variable domain sequences were modeled with the ABodyBuilder2 module of the ImmuneBuilder program and validated with QMEANDisCo (69, 70). Epitope prediction was performed through blind docking assays using ClusPro antibody mode (71). Prior the submission to ClusPro, residue charges were adjusted to pH 7.4 using PDB2PQR (72). Spike structures from 10 VOCs were recovered from the Protein Data Bank using the IDs: 6VYB (Wuhan); 8DLI (Alpha); 8DLL (Beta); 8DLO (Gamma); 7W9E (Delta); 7XO5 (Omicron BA.1); 8XZH (BA.1.1.529); 7XOA (BA.2); 7XIY (BA.3); 7XNQ (BA.4/5). The lowest-energy complex from each docking run was obtained, and the complexes were aligned in PyMOL to check for convergent binding poses (73). The interacting amino acids were identified and used as active residues in an information-driven docking step using HADDOCK2.4 and the Spike protein (PDB ID: 6VSB), with the aim of refining the antibody-antigen interface (74, 75). The results were assessed based on cluster size, buried surface area, and HADDOCK score. Moreover, binding energy was estimated using the PRODIGY webserver and protein-protein interactions were identified with PDBSum (76, 77).

A final validation step applied heated Molecular Dynamics (MD) to assess the thermostability of the HADDOCK-generated complexes, distinguishing true binding poses from decoys (78). Briefly, AmberTools19 was used to insert the complexes into a truncated octahedral box which was solvated with SPC/E water and adjusted to 0.15 M NaCl concentration. The energy of the systems was minimized. Them, they were heated to an initial temperature of 310 K and equilibrated in two subsequent steps prior to a 70 ns production run. The temperature increased to 330, 360, and 390 K at 30, 42.5, and 55 ns, respectively. All simulation steps were conducted in Amber18, and the trajectories were analyzed in *cpptraj* to evaluate the interface RMSD using the cut-off of 5 Å to identify stable poses. Three replicates of 150-ns regular MD simulations were ran using the validated complexes at the temperature of 310 K. 100 frames from the last 50 ns were used to extract the binding free energy (ΔG_bind_) of the antibody-antigen complexes through MM/GBSA analysis implemented in the MMPBSA.py application (79).

### Statistical analysis

Statistical analyses were performed using GraphPad Prism 10.0 software. Using a statistical model based on the Poisson distribution, the percentage of negative cultures obtained in ELISA assays of LDA cultures, plotted as a function of the number of cells per well, allowed estimation of the frequency of Ig-secreting B cells responsive to LPS stimulation and the frequency of Ig-secreting B cells with specific reactivity to a given antigen (18, 19, 80). Tests were chosen according to the type of variable and indicated in each result. Results with P > 0.05 were considered significant.

## Data Availability

All data produced in the present study are available upon reasonable request to the authors

## Acknowledgments

We extend our gratitude to all study participants for their time and contribution to our research. We thank the members of the Laboratório de Instrumentação Nuclear (CT - UFRJ) for their assistance with cell irradiation; the Unidade Multiusuário de Citometria - IMPG (CCS/UFRJ) team for their support with the MoFlo sorter (Dako Cytomation) and technical assistance; Ronaldo Rocha (LAGIIVIR, IMPG – UFRJ) for technical assistance. We are grateful to Dr. Garnett Kelsoe (Duke University, Durham, NC) for generously providing the NB21 feeder cells and to Victor Ramos (Laboratory of Molecular Immunology, The Rockefeller University) for sharing the human V gene database and supporting the V gene segment analysis. We thank Professor Pedro Moreno Pimentel-Coelho from the Laboratório de Neuropatologia Experimental (LINE - UFRJ). We also thank all the GIISER-Brazil members for their contributions.

## Author contributions

Conceptualization: LC, DLO, AB and AMV

Performed experiments: LC, DLO, GM, FC, DARS, YM, and AMV

Experimental support: GMA, BG, LT, BS, PC and BM

Patient enrollment: AT, OCFJr and TMPPC,

Sample processing and storage: LC, GM, DASR, BG, JE, and MB

Spike proteins production: LRC

Pseudo neutralization assays: SSF, MSC, LJC

Antibody expression and purification: CN, MCE, MNR and LFF

*In silico* analysis of antibody binding: AOA and JHMS

Intellectual contribution: OCFJr, LFF, LJC, AN, GDV, CL, AB and AMV

Analyzed data: LC, DLO, GM, FC, AOA, AN, AB and AMV

Funding acquisition: AT, MTB, LFF, GDV, CL, AB and AMV

Resources: FMBO, JE, MB

Writing—original draft: LC, DLO and AMV

Writing—review & editing: LC, DLO, AOA, FMBO, FJC, LFF, AN, GDV, CL, AB and AMV

## Data and materials availability

All data are available in the main text or the supplementary materials.

## Supplementary Materials

**Fig. S1.**
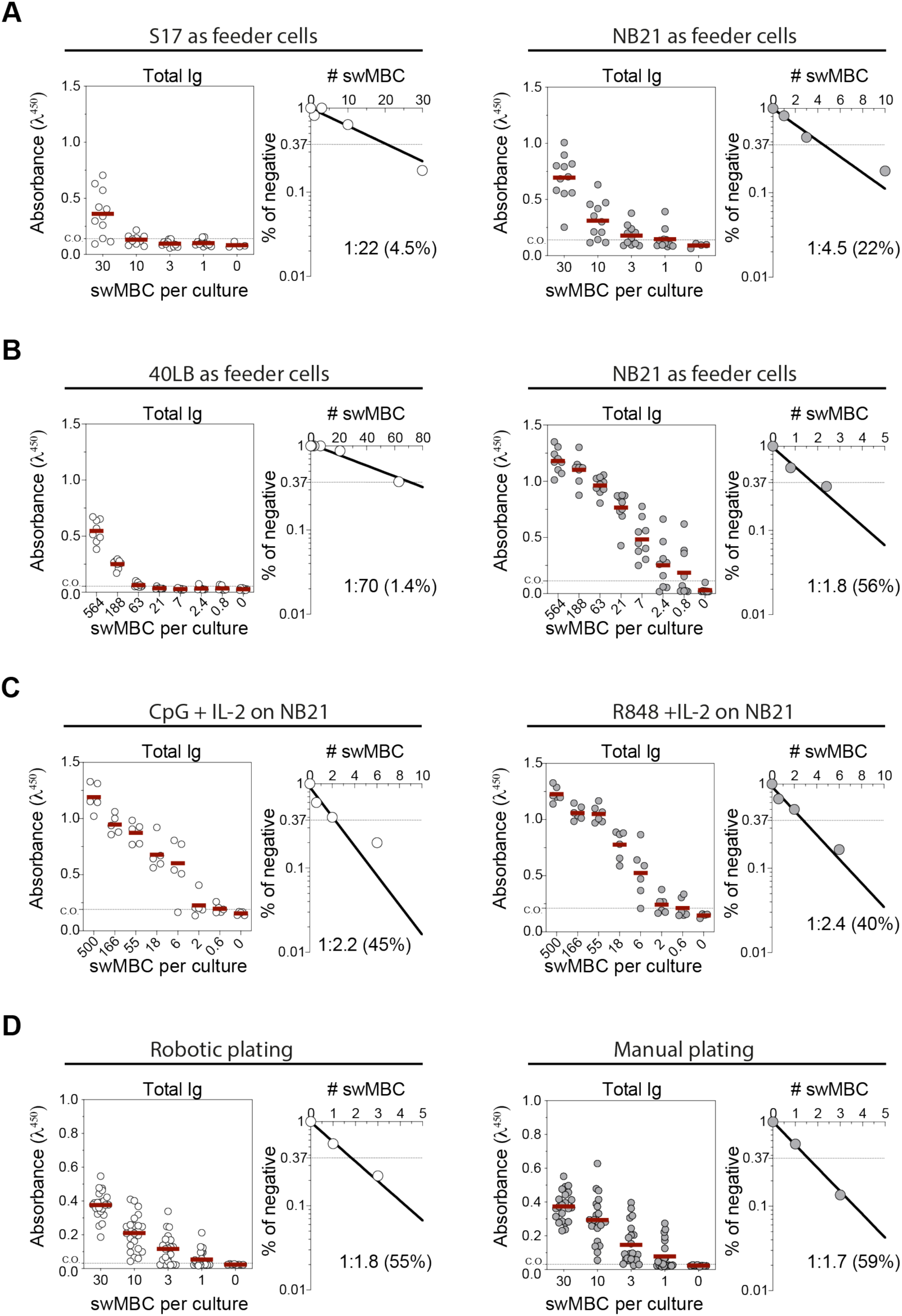
Establishing effective culture conditions for *in vitro* expansion of human memory B cells. **(A)** Frequency of responding MBCs co-cultured with S17 or NB21 feeder cells. **(B)** Frequency of responding MBCs co-cultured with 40LB or NB21 feeder cells. Decreasing number of B cells were cultured and Ig secretion was assessed by ELISA. According to the Poisson distribution, the frequency of responding cells was calculated as the inverse of the cell density at which 37% of wells lacked detectable Ig (i.e., negative cultures). **(C)** Comparison of polyclonal stimuli efficacy using CpG and R848. **(D)** Comparison of responding cell frequencies obtained by manual plating versus automated robotic plating integrated into the sorting device.

**Fig. S2.**
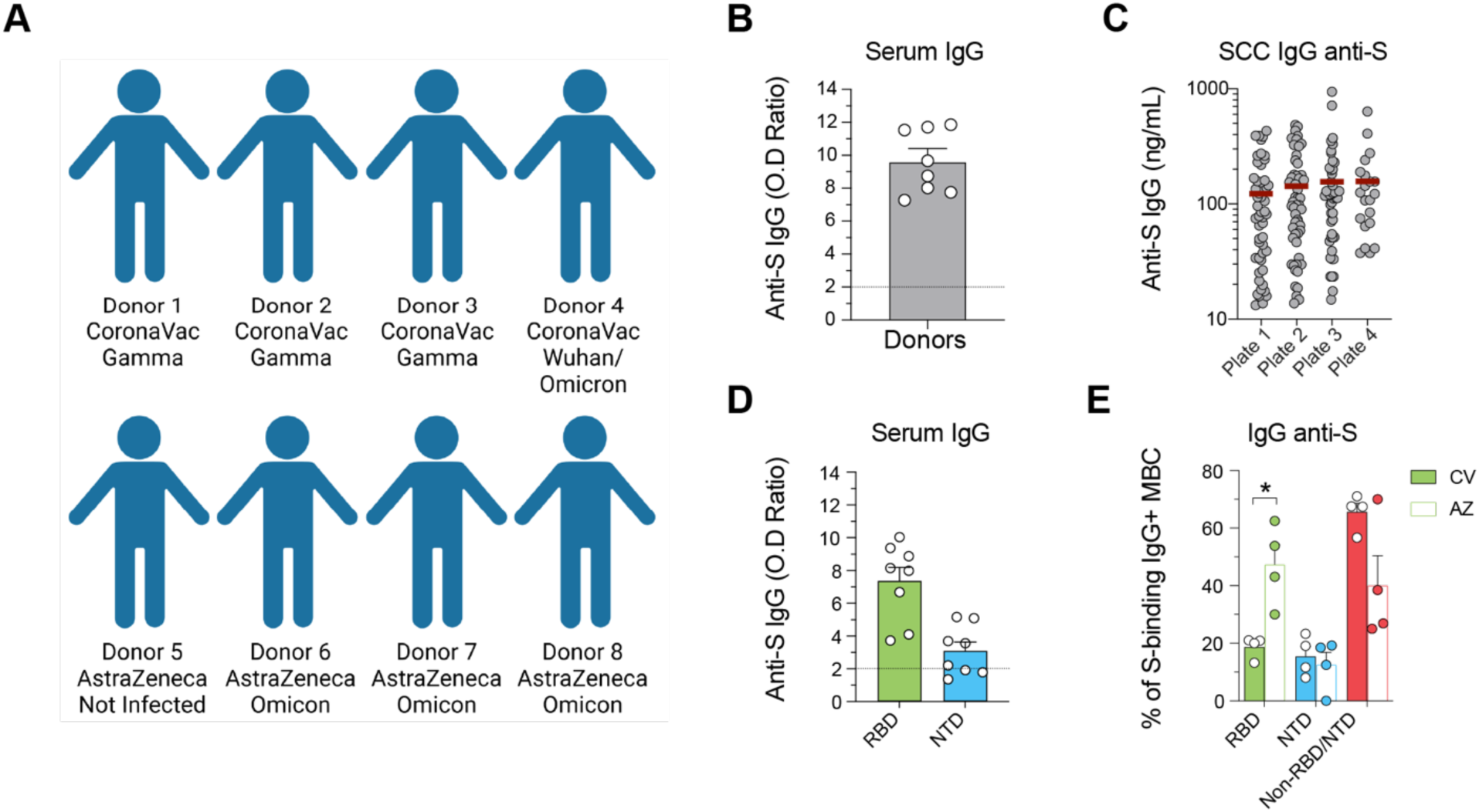
Vaccination and infection history of patients used for monoclonal antibody isolation. **(A)** Schematic representation of the vaccination and infection history for the eight donors from whom monoclonal antibodies were isolated. **(B)** OD (optical density) ratio of total IgG specific to the Spike protein in the serum of the eight donors. **(C)** Levels of anti-Spike IgG antibodies in SCC supernatants are expressed as ng/mL equivalents, quantified using a Wuhan anti-Spike human monoclonal antibody (CZmAb) as the standard. Four independent ELISA plates yielded average concentrations of 123 ng/mL, 143 ng/mL, 156 ng/mL, and 157 ng/mL of Spike-specific IgG. **(D)** OD ratio of IgG specific to the RBD and NTD regions in the serum of the eight donors. **(E)** Percentage of Spike-specific MBCs of each patient categorized by binding to the RBD, NTD, and undefined regions. Data are stratified by the type of vaccine administered to the patients: CoronaVac (CV), filled bars, or AstraZeneca (AZ), open bars. Statistical analyses were performed using the unpaired two-tailed Student’s t test. Significant difference is indicated by asterisk (*, P < 0.05; **, P < 0.01; ***, P < 0.001; ****, P < 0.0001). Error bars represent SEM.

**Fig. S3.**
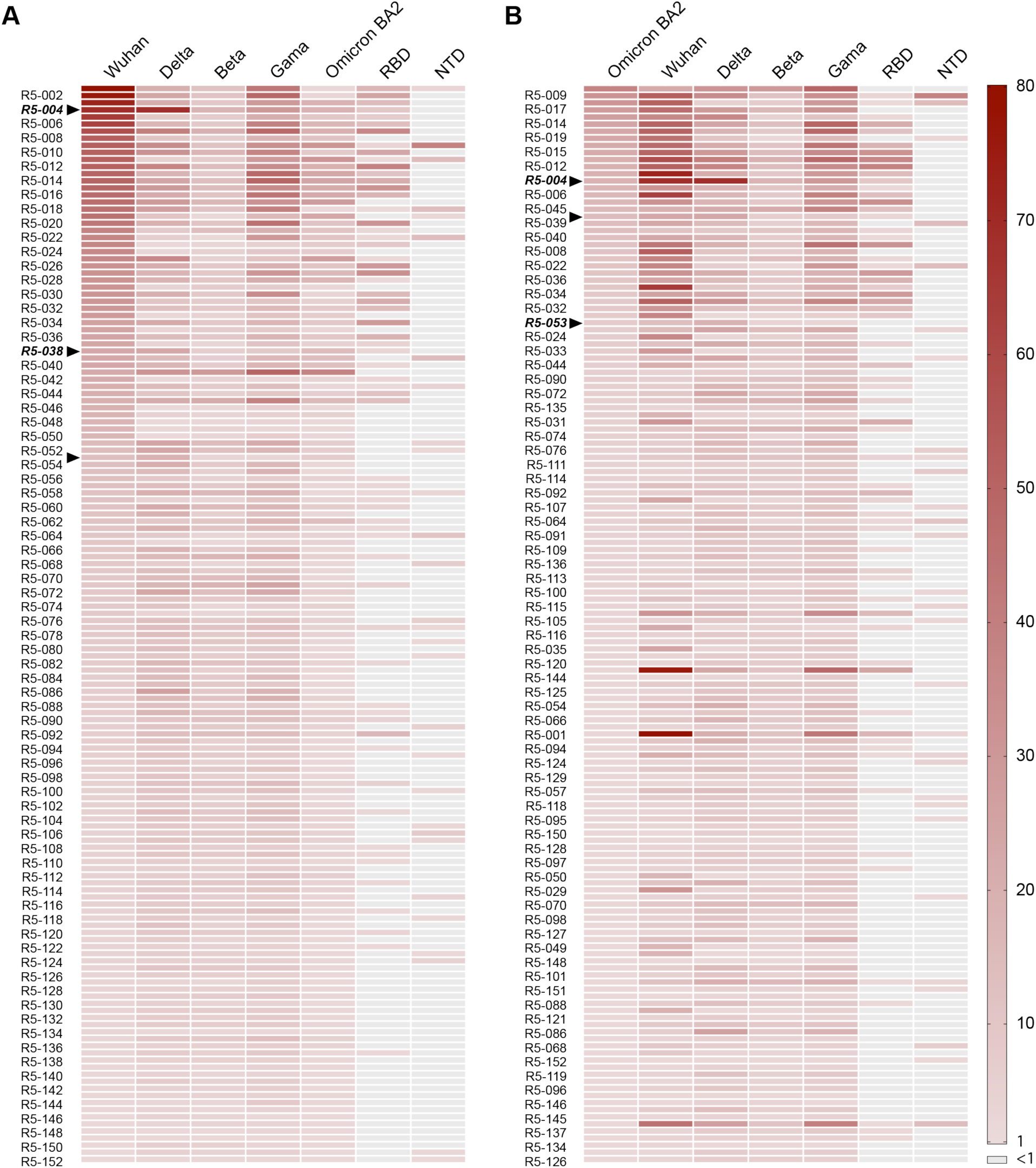
Cross-reactivity levels of mAbs evaluated by ELISA. Heatmap showing the binding intensity of monoclonal antibodies (mAbs) to variants of concern (VOCs) in ELISA assays. **(A)** R5 clones were ranked based on their OD (optical density) ratio values in the ELISA assay against Wuhan Spike and **(B)** Omicron Spike. Arrows indicate clones selected for pseudoneutralization assays.

**Fig. S4.**
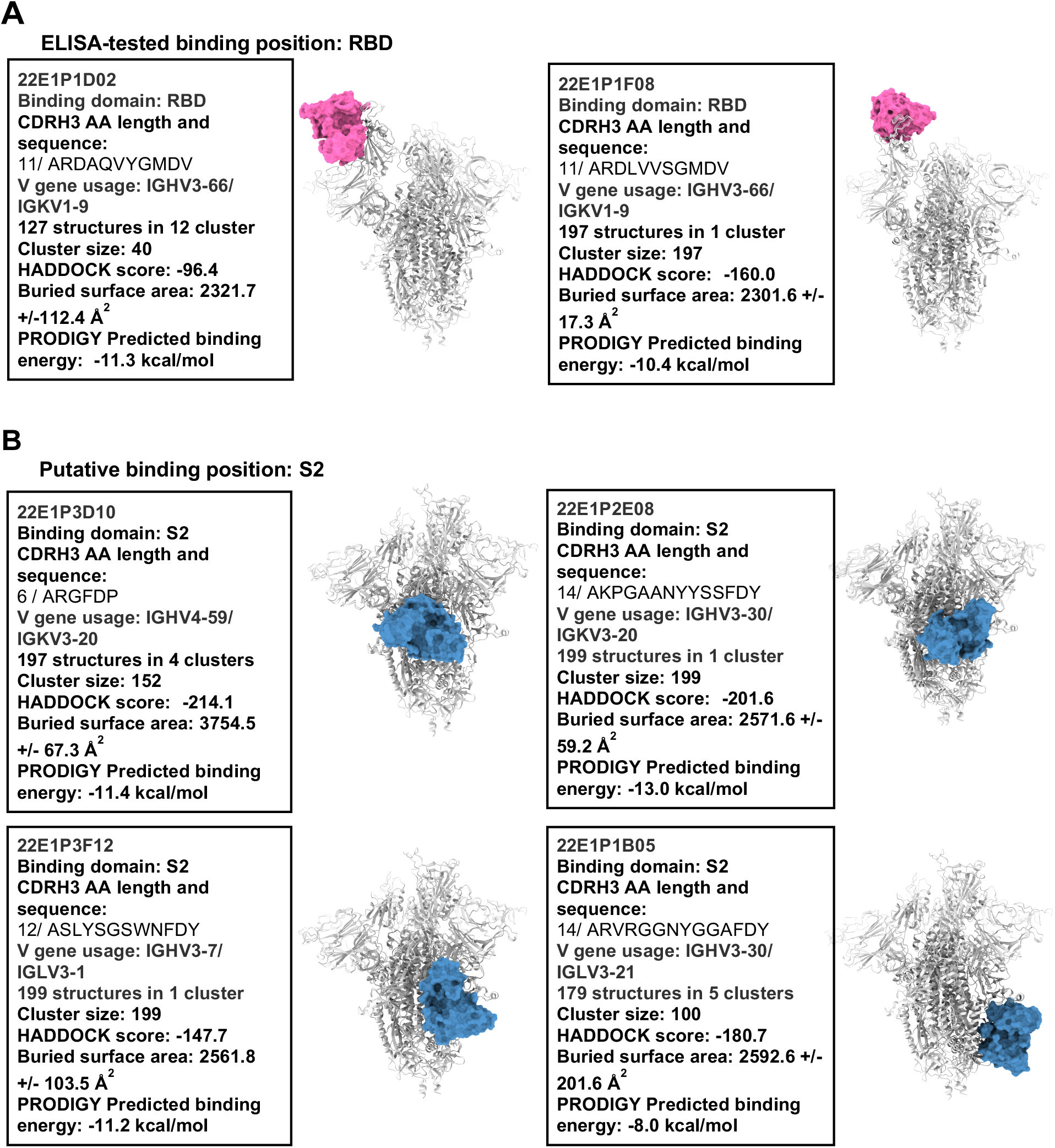
*In silico* analysis of antibody binding. **(A)** Driven docking data of the two selected RBD-binding clones. **(B)** Driven docking data of selected RBD-/NTD-, putative S2 binding clones, that shared CDRH3 sequence homology and V_H_ gene segments with clustered clones that are known to bind S2 domain.

**Fig. S5.**
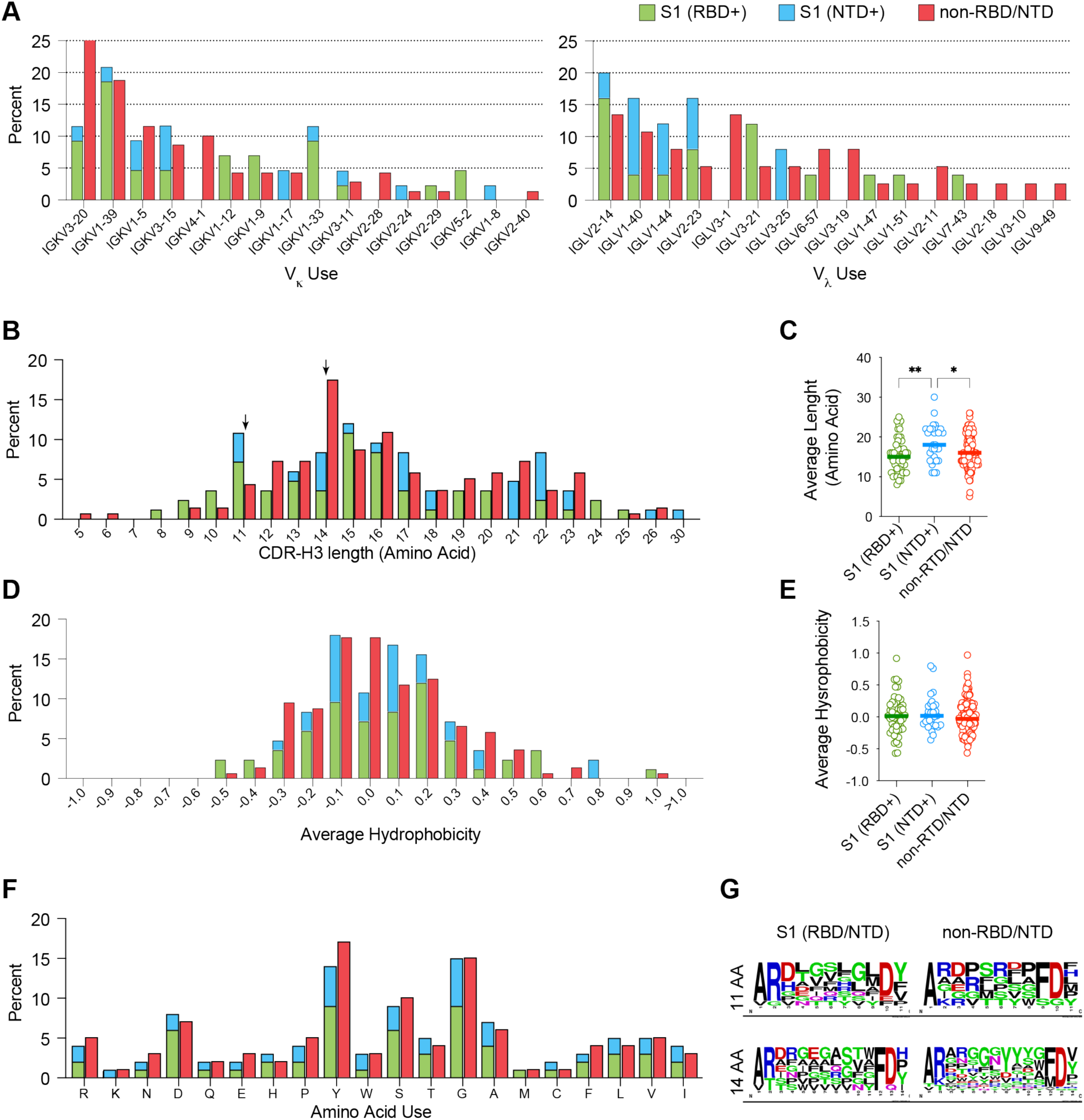
IGKV and IGLV gene usage and CDR-H3 repertoire analysis. (**A**) Distribution of CDRL3 V gene usage in κ and λ alleles for clones binding RBD, NTD, or RBD⁻/NTD⁻ Spike regions. **(B-C)** CDRH3 amino acid length distribution **(B)** and average length **(C)** for RBD, NTD or RBD-/NTD-S binding clones. Statistical analyses were performed using one-way ANOVA with Tukey’s multiple comparison test. Significant difference is indicated by asterisk (*, P < 0.05; **, P < 0.01). **(D-E)** CDR-H3 average hydrophobicity index variation **(D)** and average hydrophobicity distribution **(E)** for RBD, NTD or RBD-/NTD-S binding clones. **(F)** Amino acid usage in CDRH3 of the assessed clones. **(G)** Logo plot of amino acid usage for clones binding RBD/NTD or neither, restricted to CDR-H3 loops of 11 or 14 amino acids.

**Fig. S6.**
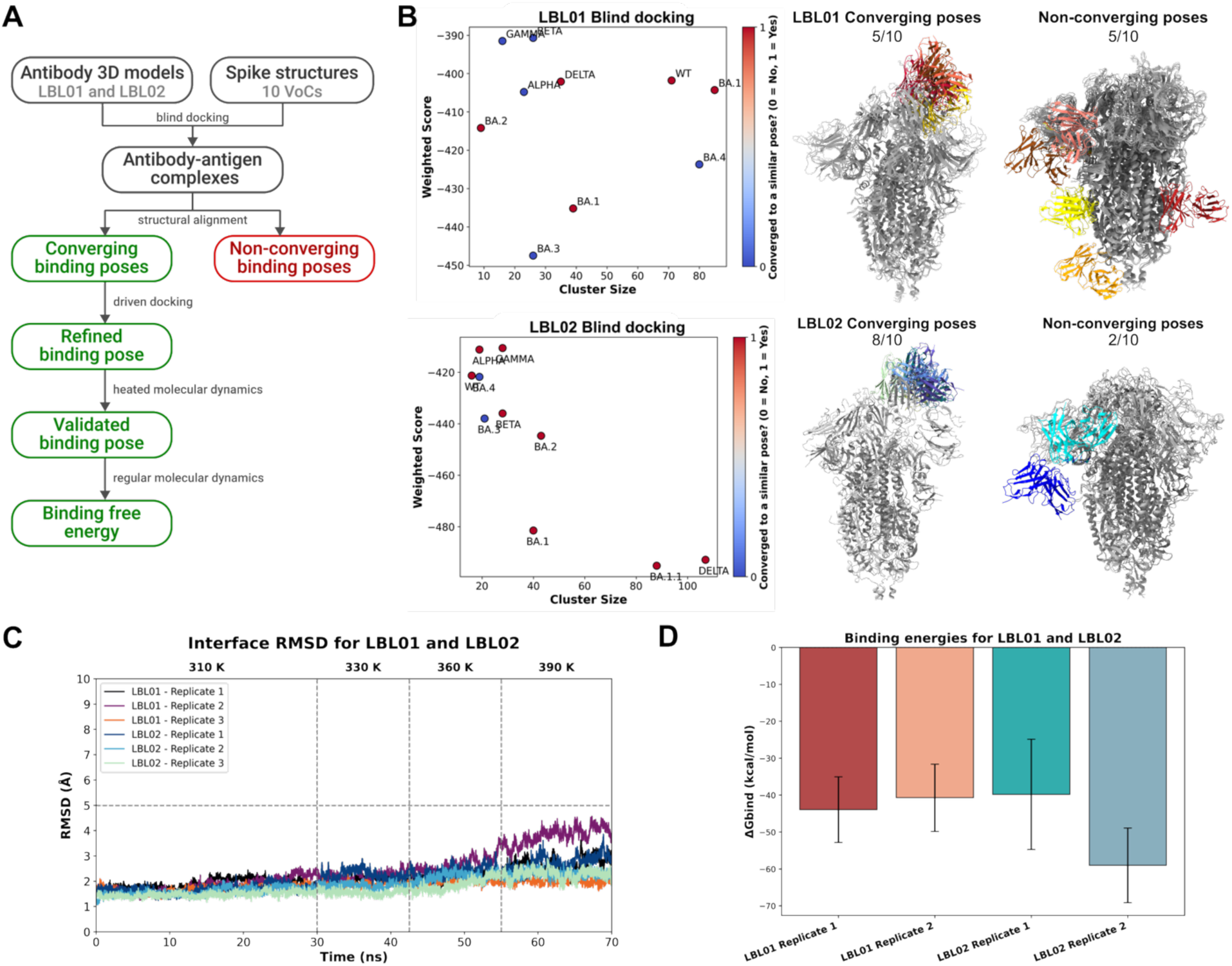
LBL01 and LBL02 docking and molecular dynamics analysis. **(A)** Computational workflow used to determine the binding poses and estimate the binding free energies of LBL01 and LBL02 with the Spike protein. **(B)** Blind docking of LBL01 and LBL02 with the Spike from 10 VOCs using ClusPro Antibody Mode compared in terms of cluster size and weighted score. Poses that converging to a similar region of Spike are shown in red, whereas the ones dispersed in other potential epitopes are in blue. Alignment of the docked complexes indicated that the converging poses bound to the RBD, suggesting a preferred binding mode. **(C)** Interface RMSD values of the antibody-RBD complexes during three independent replicates of heated MD simulations. A 5 Å cut-off was applied to identify stable binding poses, considered representative of true binding modes. **(D)** Binding free energy (ΔG_bind_) of the LBL01 and LBL02 to RBD, obtained through MM/GBSA analysis of 150 ns MD simulations. More negative values indicate a more favorable interaction.

**Table S1.**
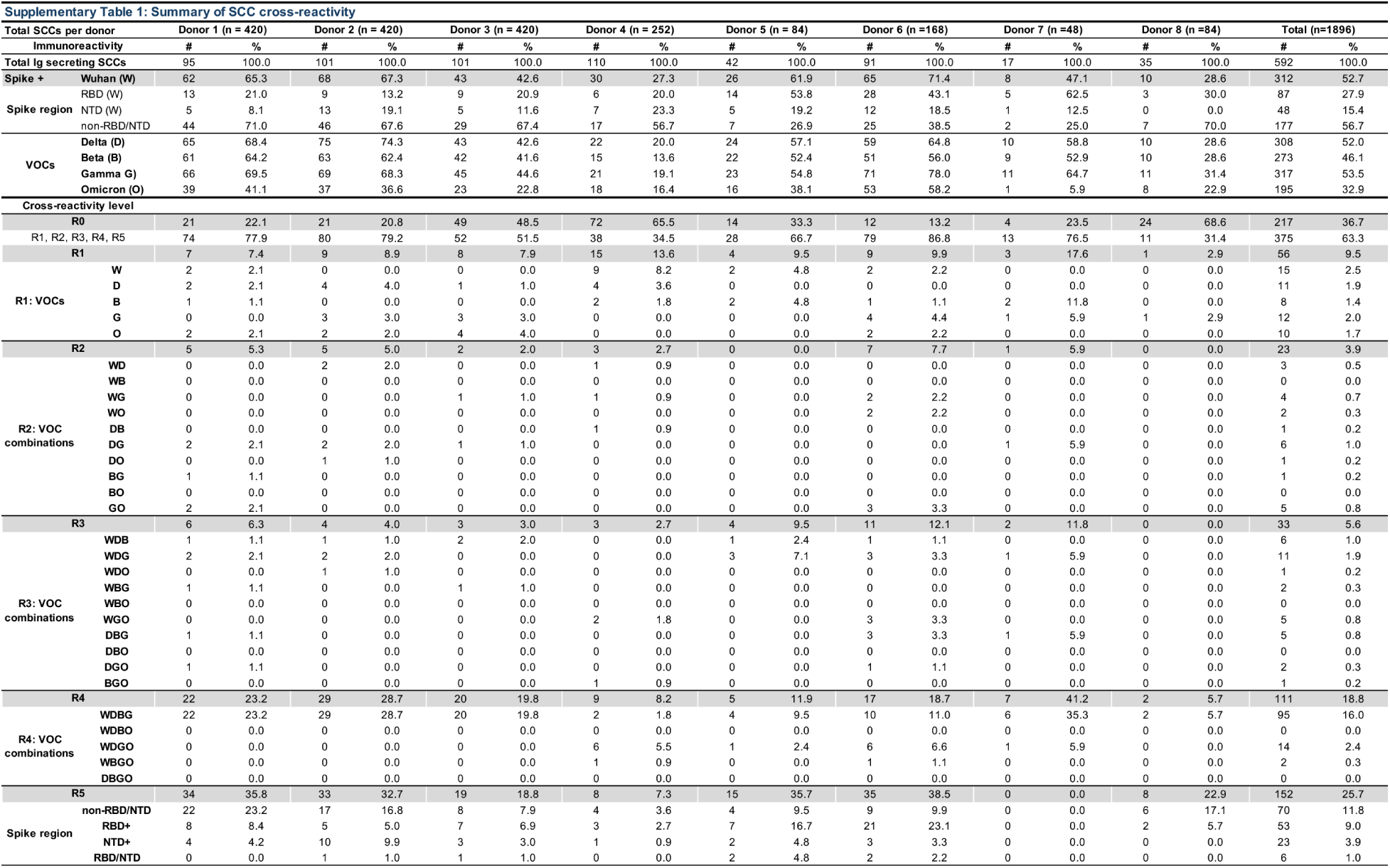
Summary of site-specific and cross-reactive single cell cloning cultures.

